# Airborne PM_2.5_ and the Emergence of 10 SARS-CoV-2 Variants The Multifaceted Influence of an Airborne Pollutant on Viral Natural Selection determining SARS-CoV-2 Evolution - An Environmental Wake-up Call or an Ecological Fallacy?

**DOI:** 10.1101/2021.06.27.21259602

**Authors:** Yves Muscat Baron

## Abstract

**Background:** Airborne particulate matter has been suggested as a co-factor for SARS-CoV-2 infection. Besides the deleterious effect this pollutant has on pulmonary immunity and the propagation of respiratory ACE-2 receptors (angiotensin converting enzyme II), the SARS-CoV-2’s point of entry, particulate matter has also been proposed as a vector for this virus’ transmission. Particulate matter may also be a marker for anthropogenic activity acting as a surrogate for increased human to human contact, increasing both transmission and the mutagenic viral load. Genes coding for SARS-CoV-2 have been detected on airborne particulate matter and its proximity to the virus, may have caused this pollutant to act as a mutagen causing the inception of SARS-CoV-2’s variants and simultaneously being genotoxic to the progenitor viruses, differentially favouring variant emergence.

Since the initial phases of the pandemic, a multitude of SARS-CoV-2 variants have been detected, but the few that survive to promulgate human infection have increased transmissibility. It also appears that there is a limited set of persistent mutations SARS-CoV-2 can produce. This set of mutations has been found in widely disparate and distant regions. This may suggest that besides intra-host mutation in an inflammatory ambience, an ubiquitous factor such as an environmental mutagen, may have resulted in convergent evolution leading to the emergence of similar variants. This paper examines a possible association in a multi-modal manner between the airborne pollutant PM_2.5_ and the emergence of ten of the most clinically and epidemiologically relevant SARS-CoV-2 variants.

**Methods:** The daily average levels of PM_2.5_ of a number of cities, where variants were detected, were obtained from the World Air Quality Index (WAQI), a real-time assessment of atmospheric pollution. PM_2.5_ levels were correlated with SARS-CoV-2 variants including Variants of Concern (VOC) or Variants of Interest (VOI). These variants included the G614 variant in Beijing, the 20A.EU1 variant in Valencia, the B.1.351 variant in South Africa, the B.1.1.7 variant in the UK, the USA variants B.1.429 in Los Angeles, B.1.2 in Louisiana and New Mexico, the B.1.526 variant found in New York, the variant B.1.1.248 in Brazil. During mid-March 2021, the B.1.617 variant first detected in October 2020, surged in Nagpur, India and the R.1 variant was detected in Kentucky U.S.A. The average daily PM_2.5_ levels were assessed, the evaluation initiating just before the occurrence of the first spike/s in this pollutant’s atmospheric concentration, till after the emergence of the variants. Where available the daily number of new cases of COVID-19 diagnosed was matched to the PM_2.5_ levels.

**Results:** There appears a common pattern of PM_2.5_ in most of the regions prior and during the emergence of the SARS-CoV-2 variants. An initial spike/s of PM_2.5_ were noted on average 50 days prior to the emergence of the variants and another smaller spike/s in PM_2.5_ were noted just before or contemporaneous with the emergence of the variant. Prior to the emergence of to the G614 variant in Beijing, the average PM_2.5_ level during its peaks was 153.4µg/m3 (SD+/-63.9) to settle to a baseline of 94.4µg/m3 (SD+/-47.8)(p<0.001). Before the appearance of the 20A.EU1 variant in Valencia, the PM_2.5_ spikes averaged at 61.3µg/m3 (SD+/-21.8) to decrease to a mean of 41.2µg/m3 (SD+/-15.5) (p<0.04). In Kent, U.K. a solitary PM_2.5_ spike averaged 82µg/m3 (SD+/-29) before the detection of the B.1.1.7 and following the PM_2.5_ spike the baseline level of this pollutant was 27.8µg/m3 (SD+/-18.0) (p<0.03). In Nelson Mandela Bay South Africa, where B.1.351 was first detected, the PM_2.5_ mean baseline level was reported as 40.4µg/m3 (SD+/-14.0), while prior to this variant’s emergence, the PM_2.5_ spike averaged 85.1µg/m3 (SD +/-17.3)(p<0.0001). In Brazil the average PM_2.5_ during its spike was 107.4µg/m3 (SD+/-34.2) before B.1.1.248 variant emerged and after the spike the baseline PM_2.5_ was 48.3µg/m3 (SD+/-18) (p<0.0001). In the USA the average PM_2.5_ peak levels prior to the emergence of the SARS-CoV-2 variants were 118µg/m3 (SD+/-28.8) in Los Angeles (baseline 66.1µg/m3 (SD+/-25.1), 75+/-27.8µg/m3 (baseline 43.3(SD+/-14.4)µg/m3 in Louisiana, 71.4+/-11.3µg/m3 (baseline 43.6(SD+/-12.4)µg/m3 New Mexico, 54.3+/-13.8µg/m3 (baseline 34.4(SD+/-11.6)µg/m3 in New York and 37.7+/-7µg/m3 (baseline 28.5 SD+/-6.8)µg/m3µg/m3 in Eastern Kentucky. All the spike patterns of PM_2.5_ levels noted in the USA were significantly higher when compared to their respective baselines (p<0.0001). Prior to the surge of the variant in India, the PM_2.5_ spike in Nagpur averaged 166.8+/-10.8µg/m3 (baseline 123.2SD+/-16.9µg/m3) (p<0.0001). In the regions where the quantity of daily new cases was available, a number of significant correlations were obtained between PM_2.5_ levels and the number of new cases of SARS-CoV-2 in most of the regions reviewed.

**Conclusion:** There appears to be an association between the levels of atmospheric PM_2.5_ and the emergence of SARS-CoV-2 variants. In most regions two groups of spike/s of PM_2.5_ were noted prior to the emergence of these variants. The first PM_2.5_ spike/s approximately 50 days before the variant’s emergence may suggest that anthropogenic activity was increased possibly reflecting augmented human to human contact, consequently increasing the viral burden of the progenitor virus. The first PM_2.5_ spike may also have made populations more susceptible to SARS-CoV-2 through the propagation of the respiratory ACE receptor. There is the potential that coronavirus-laden, PM_2.5_ induced mutagenesis in the SARS-CoV-2 genome resulted in establishing persistent variants and contemporaneously was genotoxic to the progenitor virus, expediting the latter’s disappearance. PM_2.5_ may have further diminished the pulmonary immunity inviting further viral invasion. The second spike/s prior to the emergence of variants, may suggest another anthropogenic spike in human activity. With the second spike/s in PM_2.5,_ this airborne pollutant may have acted as a viral vector encouraging variant emergence. This may have not only led to increasing viral transmission, catalysed by the preceding risk factors, but resulted in an overwhelming viral load, providing fertile ground for variant emergence. The above findings suggest that antecedent spikes in PM_2.5_ prior to variant emergence not only contributed to transmission, but also impacted the immediate viral environs which resulted in its natural selection, effecting SARS-CoV-2’s evolution.

## INTRODUCTION

The SARS-CoV-2 pandemic has reappeared in subsequent waves in the form of more transmissible and potentially more virulent variants. This increase in COVID-19 incidence has been attributed to the reversal of regional lockdowns and measures, endorsing physical distancing which were legally enforced in most countries. Following lockdown, a contemporaneous reduction in COVID-19 rates and atmospheric pollution including particulate matter (PM_2.5_) were noted only to be partially reversed once lockdown was ceased. This reversal of lockdown also involved a recrudescence of elevated levels of the pollutant particulate matter PM_2.5_.

The first study suggesting an association between PM_2.5_ and SARS-CoV-2 was noted in the United States, whereby a link between long-term exposure to particulate matter PM_2.5_ and COVID-19 related mortality was demonstrated (Wu et al 2020). A recent preprint has confirmed that PM_2.5_ was a robust variable in connection with increasing SARS-CoV-2 rates (Milicevic et al 2021). Morphological evidence also confirmed that genes coding for SARS-CoV-2 were found attached to particulate matter (Setti et al 2020a). In a large number of Chinese cities, a 2% increase in COVID-19 new cases was demonstrated with every 10μg/m3 increment in airborne PM_2.5_ (Zhu et al 2020). The deleterious effects of particulate matter on pulmonary microbial defences may encourage SARS-CoV-2 colonization of the respiratory epithelium (Domingo and Rovira, 2020), (Paital and Agrawal, 2020). Particulate matter increases the number of ACE-2 receptors, the point of host cell entry of SARS-CoV-2 (Paital and Agrawal, 2020), Sagawa et al., 2021). Particulate matter may actually act as a vector for transmission of COVID-19 infection by increasing its airborne reach surrounding the human habitat (Comunian et al. 2020) (Contini and Costabile, 2020) and also aiding deeper penetration into the respiratory tract (Qu et al., 2020).

It has been suggested that particulate matter PM_2.5_ was not only responsible for SARS-CoV-2 transmission, but may also have been involved in this virus’ evolution (Muscat Baron 2020a), (Muscat Baron 2021a). Acting as a SARS-CoV-2 vector, PM_2.5_ may have been responsible for exerting selective pressure determining the emergence of the first variant called the G614 variant (Muscat Baron 2020a), (Muscat Baron 2021a).

A recurring set of mutations, mainly E484K, N501Y and K417N suggests SARS-CoV-2 may be undergoing convergent evolution (Gupta 2021). This convergent evolution may be catalyzed by the presence of a common environmental mutagen such as the ubiquitous pollutant PM_2.5_ which has repeatedly been shown to be a robust co-factor in the SAR-CoV-2 Pandemic (Milicevic et al 2021) (Setti et al 2020a) (Muscat Baron 2020a; Muscat Baron 2020b; Muscat Baron 2020c; Muscat Baron 2020d; Muscat Baron 2020e).

Contrasting effects by PM_2.5_ on viral infectivity have been shown on two bacterophages Φ6 and ΦX174 (Groulx et al 2019). Aerosol admixture of PM_2.5_ with bacteriophages Φ6 and ΦX174 (a non-enveloped bacteriophage) showed a reduction of Φ6 infectivity but a contrastingly superior ΦX174 infectivity compared to controls (Groulx et al 2019).

This paper examines the possibility that pollution with PM_2.5_ changes augmented COVID-19 infection and had a hand in SARS-CoV-2’s natural selection, strongly suggesting that particulate matter may have acted co-factor in a multi-modal manner catalyzing the SARS-CoV-2 Pandemic through the emergence of its variants.

## METHODS

The average levels of particulate matter PM_2.5_ of a number of cities were obtained from the World Air Quality Index (WAQI). This Air Quality Index is a real-time measurement of atmospheric pollutants, including PM_2.5_ (EPA Environmental Protection Agency 2020-2021). The daily average PM_2.5_ levels were assessed prior to a prominent spike in PM_2.5_ till beyond the emergence of the SARS-CoV-2 variants in each region. During 2020, the regions noted to have COVID-19 variants included Beijing, Valencia, Nelson Mandela Bay in South Africa, Bexely (U.K.), Los Angeles, New York, Louisiana, and New Mexico in the USA and Sao Paolo in Brazil. Recently in mid-March 2021, two variants emerged in Nagpur, in the Maharashtra region of India and in Eastern Kentucky U.S.A respectively. In Beijing, the period between January 20^th^ and mid-February 2020 was chosen when the G614 variant (Chart 1. and 2.) with the G614 clade appears to have been detected (Korber et al 2020). In Valencia the first peak of PM_2.5_ (Chart 3. and 4.) was noted around the 11^th^ of April and the 20A.EU1 variant appears to have been detected at the end of May 2020 (Hodcroft et al 2020). The PM_2.5_ in Bexely, U.K. peaked on the 12^th^ August 2020, 40 days before the B.1.17 variant was detected on the 20^th^ of September (Chart 5. and 6.). In Nelson Mandela Bay the origins of the B.1.351 variant in South Africa were noted in early October, 60 days after the PM_2.5_ peak on the 19^th^ July 2020 (Chart 7.). In Los Angeles the first case of the B.1.429 variant was noted during a solitary PM_2.5_ spike on the 6th of July which was followed by another two wider spikes in mid-September and the beginning of October after which a surge in COVID-19 cases was noted in November (Chart 8. and 9.). The B.1.526 variant was detected in the in New York at the end November, 15 days after a spike in PM_2.5_. The surge in the B.1.526 variant followed in mid-December contemporaneous with a PM_2.5_ spike (Chart 10. and 11.). In Louisiana and New Mexico, the B.1.2 variant followed a similar pattern with three spikes in PM_2.5_, the first and most prominent 45 days before the variant was detected in September, the second spike at detection in November and the third smaller spike in early December when COVID-19 cases surged (Chart 12, 13, 14 and 15). In Sao Paolo Brazil two peaks of PM_2.5_ were noted during September to be followed by the emergence of the B.1.1.248 variant in early December (Chart 17).

Two variants emerged in mid-March 2021, one occurring in Nagpur India (Chart 18. and 19.) and the second in Eastern Kentucky, USA(Chart 16). Prior to the B.1.617 variant in India a wide peak of PM_2.5_ was noted during the first three weeks of January and 50 days later, during another smaller peak, the B.1.617 variant emerged in mid-March 2021. After a series of PM_2.5_ peaks in February in Kentucky the R.1 variant was detected 45 days later, contemporaneous with another larger PM_2.5_ spike in mid-March 2021.

The viral samples first determining the G614 variant in China were obtained from Beijing (Korber et al. 2020). This does necessarily mean that the variant first emerged in Beijing, however for lack of further evidence the PM_2.5_ in Beijing was utilized in this study. It may well be that with the sudden rise in cases the G614 variant first emerged in Wuhan. The paper by Hodcroft et al. point to the North-East of Spain as being the origin of the 20A.EU1, and following a superspreader event in Bergamo, the PM_2.5_ levels from Valencia were assessed. The B.1.1.7 appears to have originated in Kent, U.K. and the PM_2.5_ levels in Bexely were utilized in this study. The B.1.351 variant in South Africa has been suggested to have possibly originated in Nelson Mandela Bay where although PM_2.5_ levels are available, the daily new case counts could not be collated from the East Cape. In Los Angeles the first case of the B.1.429 variant was noted during a solitary PM_2.5_ spike while the B.1.526 variant was detected in the Washington Heights in New York. In Louisiana and New Mexico, where the B.1.2 variant was first detected, the PM_2.5_ levels of these States’ capital cities were assessed. The PM_2.5_ levels in East Kentucky were used in this study as this is where the R.1 variant is reported to have been first detected. The PM_2.5_ levels of Sao Paolo were assessed as this was pollution data available from Brazil, although the B.1.1.248 variant appears to have been first detected in Manaus which however did not have any PM_2.5_ levels available on WAQI. The pattern of elevated levels of particulate matter pollution in Brazil appears to perennially occur in October due to anthropogenic and natural factors. The B.1.617 variant appears to have been first detected in India in November 2020, but its emergence as a VOC was noted in Nagpur in Mid-March 2021. As may be noted in the absence of solid evidence, in some cities certain assumptions had to be taken to decide the site from which PM_2.5_ levels had to be assessed.

The data was analysed for normality and all the data were found to be nonparametric. The Mann Whitney U test was applied for comparing nonparametric variables of both groups of cities and the Spearman Rank test was applied for nonparametric correlations.

## RESULTS

There appears a common pattern of PM_2.5_ in most of the regions assessed prior and during the emergence of the COVID-19 variants. An initial spike/s of PM_2.5_ was noted on average 50 days prior to the surge of the variants and another smaller spike/s in PM_2.5_ was noted just before or contemporaneous with the emergence of the variant (Charts 1-18). Prior to the emergence of the G614 variant in Beijing, the average PM_2.5_ level during its initial spike was 153.4µg/m3 (SD+/-63.9) to settle to 94.4µg/m3 (SD+/-47.8)(p<0.001)(Chart 1.). With the data available the number of new cases of COVID-19 in Beijing decreased with diminishing PM_2.5_ levels (Chart 2). Before the appearance of the 20A.EU1 variant in Valencia the PM_2.5_ three spikes averaged at 61.3µg/m3 (SD+/-21.8) to decrease to a mean of 41.2µg/m3 (SD+/-15.5) (p<0.04)(Chart 3.). Similar but to a lesser extent COVID-19 new cases in Valencia decreased with decreasing PM_2.5_ levels (Chart 4.). In Kent, U.K. the single initial PM_2.5_ spike averaged 82µg/m3 (SD+/-29) before the detection of the B.1.1.7 when the PM_2.5_ averaged 27.8µg/m3 (SD+/-18) (p<0.03) (Chart 5.). As regards COVID-19 cases in Kent there was a weak correlation with PM_2.5_ levels (Chart 6). In Nelson Mandela Bay where B.1.351 was first detected, the PM_2.5_ mean level was reported as 40.4µg/m3 (SD+/-14) while prior to this variant’s emergence, the PM_2.5_ spike averaged 85.1µg/m3 (SD +/-17.3)(p<0.0001)(Chart 7.). In Sao Paolo Brazil the average PM_2.5_ during its peak was 107.4µg/m3 (SD+/-34.2) before B.1.1.248 variant emerged when the baseline PM_2.5_ was 48.3(SD+/-18) (p<0.0001)(Chart 17.). In the USA the average PM_2.5_ spike levels prior to the emergence of the SARS-CoV-2 variants were 118(SD+/-28.8)µg/m3 in Los Angeles (baseline 66.1(SD+/-25.1)µg/m3, 75+/-27.8(baseline 43.3(SD+/-14.4)µg/m3 in Louisiana, 71.4+/-11.3µg/m3(baseline 43.6(SD+/-12.4)µg/m3 New Mexico, 54.3+/-13.8)µg/m3 (baseline 34.4(SD+/-11.6) in New York and 37.7+/-7(baseline 28.5(SD+/-6.8)µg/m3 in Eastern Kentucky, all of which were significant (p<0.0001) (Charts 8 – 16). Prior to the emergence of the variant in India, the PM_2.5_ peak in Nagpur averaged 166.8+/-10.8µg/m3 (baseline 123.2SD+/-16.9µg/m3)(p<0.0001) (Charts 18.). There was no correlation between COVID-19 new cases and PM_2.5_ levels (Chart. 19.) for the Indian variant.

Where available for assessment a number of variables correlations resulted between new cases SARS-CoV-2 and PM_2.5_ levels. In Beijing the strongest correlation between PM_2.5_ concentrations and the number of new cases was noted (R=0.53 (p<0.001). A weaker correlation between PM_2.5_ concentrations and the number of new cases was obtained in Valencia (R=0.25 p<0.05). No significant correlations between PM_2.5_ concentrations and the number of new cases in Bexely U.K, Louisiana and New York, USA. Negative significant correlations resulted in Los Angeles (R=-0.36 p<0.001) and Mew Mexico (R=-0.35 p<0.0001). No correlation between COVID-19 new cases and PM_2.5_ levels, however it must be mentioned that the new cases were retrieved from the Maharashtra province whereas the PM_2.5_ levels were obtained from one of its cities, Nagpur the putative origin of the Delta variant (Indian variant).

## DISCUSSION

This paper indicates that a common pattern appears to occur in relation to PM_2.5_ levels in most of the locations where SARS-CoV-2 variants emerged. Approximately 50 days prior to the emergence of the ten most persistent SARS-CoV-2 variants, a spike/s in the atmospheric levels of the airborne pollutant PM_2.5_ was noted for most of the variants that appear to have continued to promulgate human infection. Just before or contemporaneous with the variant surge, another smaller spike/s in PM_2.5_ was also noted for most of the SARS-CoV-2 variants.

Similar to this study, a lag phase in the emergence of influenza infection has been noted following exposure to particulate matter PM_2.5_. A study in Montana showed that the rate of influenza in winter increased following wildfires in summer. Elevated daily mean PM_2.5_ concentrations during the summer wildfire season positively correlated with increased rates of influenza in the following winter. With every 1 μg/m3 increase in average daily summer PM2.5, two analyses indicated a 16% and 22% increase in influenza rates respectively (Langruth et al 2020).

### Sources of PM_2.5_

The sources of PM_2.5_ very much depend on the characteristics of the regions’ activities. In the industrial regions the carbonaceous sources of PM_2.5_ are coal combustion, vehicles’ gasoline and diesel exhaust (Zhang et al 2014). A prime example is the elevated levels of PM_2.5_ in China due to the daily combustion of 80,000 tonnes of coal (Ghosh 2020). Wuhan, the site thought to be SARS-CoV-2’s origin, is considered the Chinese hub of transportation earning the moniker as the “Chicago of China”. Residential and commercial areas have similar sources of PM_2.5_ in the form of gasoline and diesel exhausts and biomass combustion due to cooking and heating.

In a trans-seasonal study done in Amsterdam and Helsinki, the sources of indoor and outdoor PM_2.5_ were very similar. In a sizable proportion (41%), the major sources of PM_2.5_ were secondary pollution, whereby primary pollutants interacted with atmospheric molecules. The remaining 59% of PM_2.5_ sources were motor vehicles’ exhaust, calcium-rich particles, biomass burning, soil and road dust, and marine aerosols. A similar pattern was noted for median personal, indoor, and outdoor PM_2.5_ concentrations, whereby in Amsterdam the levels of this pollutant were 13.6µg/m3, 13.6µg/m3, and 16.5µg/m3 respectively, while in Helsinki the PM_2.5_ concentrations were 9.2µg/m3, 9.2µg/m3, and 11.1µg/m3. In both Amsterdam and Helsinki, the personal and indoor PM_2.5_ concentrations highly correlated with outdoor concentrations (median R= 0.7-0.8) (Brunekreef et al 2005).

Specific habitats have their own species of PM_2.5_ depending on the activities carried out in these areas. In a study of schools in Barcelona, on average 47% of indoor PM_2.5_ was due to soil particles (13%) and organic sources (34%) and calcium-rich particles from chalk and building deterioration. The remaining 53% of PM_2.5_ in schools was derived from emissions from outdoor sources including week-day traffic (Amato et al 2014). Significant pollution with particulate matter may also occur emanating from natural causes such as wildfires (Meo et al. 2020). Of particular relevance especially in the U.S.A, in regions where there are perennially very low levels of PM_2.5,_ this pollutant suddenly shoots up are due wildfires. This aspect is of significance because, whereas most by-products of wildfires are rapidly transformed while suspended in the atmosphere, PM_2.5_ remains unchanged for several days (Rodgers et al 2020).

### PM_2.5_ and SARS-CoV-2 related infection and mortality

The relevance of particulate matter PM_2.5_ and PM_10_ during the pandemic became evident when epidemiological studies demonstrated that a large proportion of sporadic cases of COVID-19 could not be explained through direct human to human contact. Epidemiological studies indicated that aerosol and droplet contact could not explain these sporadic cases and the regional differences in the transmission of SARS-CoV-2 (Cai et al. 2020).

Long-term exposure to PM_2.5_ was associated with increased the SARS-CoV-2 related infection and mortality rates. A study in the USA showed that an increment of just 1 μg/m3 in PM_2.5_ exposure, correlated with an 8% augmentation in the COVID-19 mortality rate (Wu et al 2020). Another study in the USA demonstrated an increase of only 1μg/m3 in PM_2.5_ exposure was associated with a 13% increase in COVID-19 related mortality rate (Correa-Agudelo et al 2020).

In a large Chinese study of 120 cities, an increase of 10μg/m3 in PM_2.5_ was linked to with a 2.24% (95% CI: 1.02 - 3.46) rise in the daily new cases of COVID-19 respectively (Zhu et al 2020). Another study in China showed that a 10μg/m3 increment in atmospheric PM_2.5_ increased the SARS-CoV-2 case-fatality ratio by 0.24% (0.01%-0.48%) (Yao et al. 2020).

In the Lombardy region of Italy, PM_2.5_ levels as high as 38.31μg/m3 have been recorded (WHO acceptable upper limit 25μg/m3) (Comunian et al 2020). Atmospheric concentrations of PM_2.5_ in Northern Italy correlated with SARS-CoV-2 infection incidence (R = 0.67, p < 0.0001), the COVID-19 related death rate (R = 0.65, p < 0.0001) and the case-fatality rate (R = 0.7, p < 0.0001) (Comunian et al. 2020).

An international study from 63 countries over five continents similarly demonstrated a connection between PM_2.5_ and COVID-19 cases. This study showed that a 10 μg/m3 increase of PM_2.5_ pollution level was associated with 8.1% (95% CI 5.4% - 10.5%) increase in the number of COVID-19 cases during the 14 day period of assessment (Solimini et al 2021).

### PM_2.5_ and the Emergence of the G614 Variant in China

The G614 variant was the first prominent variant that appears to have originated in China early in 2020. Prior to the emergence of the G614 variant in Beijing, the average PM_2.5_ level during its peak was 153.4µg/m3 (SD+/-63.9) and later settled to 94.4µg/m3 (SD+/-47.8)(p<0.001) when the G614 variant surge occurred (Chart 1.). In Beijing the downward trend of the number of daily new cases mirrored that of the PM_2.5_ levels, however it must be mentioned that the number of new cases available were at latter part of the bell-shaped curve.

Interestingly the PM_2.5_ levels in Wuhan preceded those of Beijing by a month (Muscat Baron 2020b). This may suggest that the G614 mutation may have occurred in Wuhan with the elevated PM_2.5_ levels having “primed” the population as an ideal reservoir. The population in Wuhan may have acted as the ideal reservoir due the presence of a high viral load of the progenitor Wuhan 1, with a PM_2.5_ weakened pulmonary immunity, a PM_2.5_-induced increase in the respiratory viral point of entry (ACE receptor) in the presence of a multitude of people crammed together in constrained PM_2.5_-replete places such as train platforms or restricted spaces indoors during the Chunyun Spring Festival (Muscat Baron 2020d).

The G614 variant may have replaced the original Wuhan 1 during the mass movements of the Chinese population attending the Chunyun Spring festival which is held 15 days before the New Year and lasts a total of 40 days. The Chunyun festival is the largest mass movement occurring on the globe, involving nearly 400,000,000 individuals and over 2.9 billion journeys are made during the internal migration. A study between January 6th and February 6th, 2020 indicated that the diagnosis of COVID-19 was more likely within 11–12□days after people moved from Wuhan to 16 nearby cities in the Hubei Province. Following the spike in COVID-19 diagnoses, the number of cases declined after the implementation of cities’ lockdown (Jiang and Luo 2020). This mass movement of the Chinese population during Chunyun Spring festival overlapped with the PM_2.5_ peaks in Beijing noted in this study. Thereafter the incidence of COVID-19 in China dropped dramatically possibly due to strict social distancing, face protection and a possible innate immunity of the Chinese populations towards the G614 variant (Muscat Baron 2021b).

The G614 variant which dominated and persisted throughout the SARS-CoV-2 genomic variant landscape involved a mutation whereby an amino acid alteration in the viral spike protein gene occurred at D614G position. This mutation involved a single genomic modification with the replacement of aspartic acid by glycine at the amino acid G614 position of the spike protein. In vitro, the G614 mutation has demonstrated increased cellular infectivity, however this does not seem to be consistently the case in vivo (Korber et al 2020).

Genomic alteration at the G614 position appears to have led to a modification in the phenotypic configuration and functionality of the spike protein peptide. The original Wuhan 1 coronavirus had its spike protein’s three peptides aligned in a “closed” configuration whereas the G614 variant’s spike protein peptides are consistently found in an “open” orientation (Wrapp et al 2020).

SARS-CoV-2 adherence to the respiratory angiotensin II receptors (ACE) receptors on the pneumocyte II and goblet cells depends on spike protein’s trimeric peptide configuration. The spike protein adheres to respiratory cells’ ACE receptors if at least two of its three peptides are positioned in an “open orientation” (Yurkovetskiy et al 2020). This may also be the case impacting SARS-CoV-2 adherence to particulate matter if the latter acts as its vector.

G614 variant’s different spike protein tripeptide orientation may determine significant changes to the functionality of the receptor binding domain. Molecular dynamic computer simulations have demonstrated a variety of components, all of which may singly or in concert determine the adherence potential of the receptor binding domain. These simulations demonstrated a complex network of salt bridges, hydrophobic sites, hydrogen bonding and electrostatic interactions between the receptor-binding domain of the SARS-CoV-2 Spike protein and the angiotensin II receptor (Taka et al 2020).

### PM_2.5_ levels and the Emergence of the 20A.EU1 Variant in Spain

Three spikes in PM_2.5_ in Valencia were noted before the appearance of the 20A.EU1 variant. The PM_2.5_ spikes’ averaged 61.3µg/m3 (SD+/-21.8) to decrease to a mean of 41.2µg/m3 (SD+/-15.5) (p<0.04) when the 20A.EU1 variant emerged. Similar to the pattern in China, the trend of the number of new cases of COVID-19 mirrored that of the PM_2.5_ levels in Valencia. These spikes in PM_2.5_ overlapped with the population load in Spain following the tourist influx after travel restrictions were relaxed during May-June 2020.

A research group in Basle detected a persistent mutant designated as 20A.EU1 of SARS-CoV-2 which has spread specifically in the European Continent (Hodcroft et al 2020). The initial stages of the variant appear to have originated in the North East region of Spain. Two outbreaks of infection with this variant were detected in farmers coming from the provinces of Aragon and Catalonia in late June 2020. The 20A.EU1 variant may have originally infected the Aragonese and Catalan farmers who possibly transmitted the virus to the mink population in the North Eastern regions of Spain. Similar to humans, where population density is a risk factor for high COVID-19 infection rates, the mass confinement of minks led to widespread infection of the caged animals. This undoubtedly led to a high R_o_ factor in the confined animal population due to the exponential infection rate presenting fertile ground for the inception of variants (Muscat Baron 2021c). High reproduction rates are a prerequisite for the occurrence of mutations which eventually may flourish adapting to natural selective pressures (Domingo et al. 2016).

Spain following Italy, was one of the first European countries to have succumbed to the COVID-19 pandemic. The first Italian residents noted to have contracted SARS-CoV-2 infection were in a small town near to Milan on the 21^st^ of February (Sanfelici et al 2020). The suggestion is that a superspreader event occurred when a well attended (50,000) football match between the Spanish team of Valencia and the Italian team of Atalanta was played in the stadium of Bergamo on the 19^th^ February 2020. Unknowingly early in February, Bergamo was already the focus for seeding COVID-19 throughout the Lombardy region of Northern Italy. Following Italy, not unexpectedly, coronavirus made its appearance in Valencia and soon after the rest of Spain was plunged into lockdown as the pandemic engulfed the whole nation leading to high mortality rates in the elderly and vulnerable individuals (Mas Romero et al 2020).

Following high mortality rates, lockdown was enforced in mid-March in both Italy and Spain (Sanfelici et al 2020) (Tobías et al 2020). Social distancing succeeded in reducing the R_o_ in May, encouraging diminution of restrictions in June. Spain reopened its borders to most European tourists on the June 21 a week later than most of the European Union member states. With travel restrictions relaxed, tourists in their thousands including those from the U.K., crowded on to the Spanish coastal resorts. The holiday mood may have also relaxed the restriction on social distancing and any semblance of face protection of sun-seeking tourists. Consequently the 20A.EU1 variant spread throughout the Spanish peninsula and later the European Continent in particular the U.K. Due the paucity of transatlantic travel, the 20A.EU1 variant was not detected in the Americas.

### PM_2.5_ and the Emergence of the B.1.1.7 Variant in the U.K

Following the 20A.EU1 variant spread throughout the Spanish peninsula, Hodcroft et al. indicated that this variant spread throughout Europe. The peak of new cases of the 20A.EU1 variant was achieved in Spain in mid-August when this variant had just been detected in Kent U.K. Subsequently one month later in mid-September this variant increased exponentially in the South East of England, to soon become the predominant variant in the UK. The B.1.1.7 variant and the 20A.EU1 variant appear to evolve from the 20A progenitor clade (Lauring and Hodcroft 2021). Alternatively, possibly under the influence of an environmental factor, the 20A.EU1 variant may have reversed its mutation to the progenitor which went on the produce the B.1.1.7 variant.

Two peaks of PM_2.5_ were noted prior to the emergence of the B.1.1.7 variant. In mid-August the first spike in PM_2.5_ was noted in Kent U.K., 50 days before the emergence of the B.1.1.7 variant. A second peak in PM_2.5_ occurred just before the detection of the B.1.1.7 variant. The PM_2.5_ peaks in the South East of England averaged 82µg/m3 (SD+/-29) before the detection of the B.1.1.7 and later decreased to a mean of 27.8µg/m3 (SD+/-18) (p<0.03). This rise in PM_2.5_ possibly coincided with increased exposure to motor vehicle exhaust, as the Kent area is a busy thoroughfare during this period. Elevated levels as high as 77.2µg/m3 in Upper Stone Street in Maidstone have been detected in central Kent (Friends of the Earth).

The number of new cases of the 20A.EU1 variant in Kent coincided with the elevated levels of PM_2.5_ in this region. Having gained a foothold in September, the B.1.1.7 variant continued to slowly rise throughout October, until it exponentially increased in mid-November emerging as the dominant variant in the U.K. B.1.1.7 appears to be resistant to monoclonal antibody neutralization to the N-terminal domain of spike protein and moderately resistant a few monoclonal antibodies to the receptor-binding domain (Wang et al 2020).

### PM_2.5_ and the Emergence of the B.1.351 variant in South Africa

The B.1.351 variant was first detected in Nelson Mandela Bay, South Africa in early October 2020. The PM_2.5_ mean level in Nelson Mandela Bay during this period was reported as 40.4µg/m3 (SD+/-14) while approximately 60 days prior to this variant’s emergence, a PM_2.5_ spike averaging 85.1µg/m3 (SD +/-17.3)(p<0.0001) was noted.

The East Cape of South Africa has been hit by a 6 year drought. In July 2020 the water reservoir in Nelson Mandela Bay was down to 18% of its capacity due to the paucity of rainfall. Severe drought has a significant impact on atmospheric pollution and acts as one of largest causes of airborne PM_2.5_. PM_2.5_ aerosol concentration in the Owens Lake area in California’s southwest increases abruptly from less than 5□µg/m3 to 25□µg/m3 during the drought period (Borlina et al 2017).

The B.1.351 is considered a variant of concern as it possesses the E484K mutation. The E484K mutation appears to confer some resistance to antibodies against the SARS-CoV-2 spike protein. The B.1.351 variant appears not only refractory to neutralization by most N-terminal domain monoclonal antibodies but also by multiple individual monoclonal antibodies to the receptor-binding motif on receptor binding domain due to an E484K mutation (Wang et al 2021).

### PM_2.5_ and the Emergence of 5 Variants in the USA

Five persistent SARS-CoV-2 variants were detected in the USA. Early in May 2021 the USA variants B.1.429 and B.1.427 in Los Angeles were considered viruses of concern (VOC) while the B.1.526 and B.1.525 variants found in the Washington Heights were considered viruses of interest (VOI) (CDC 2021). Prior to May, the B.1.2 variant in Louisiana and New Mexico and the R.1. variant in Eastern Kentucky were also detected with the former soon displaced by the B.1.1.7 variant and the latter causing an outbreak in a Nursing home facility (CDC 2021).

On the 6^th^ of July the B.1.429 variant was detected in Los Angeles coinciding with a solitary peak in PM_2.5_ (Chart 8.). This peak also coincided with the use of a multitude of fireworks in the Angeles area. In Los Angeles the average PM_2.5_ peak levels prior to the emergence of the SARS-CoV-2 variants was 118(SD+/-28.8)µg/m3 in Los Angeles while its baseline was 66.1(SD+/-25.1)µg/m3. Two peaks of COVID-19 cases in Los Angeles occurred approximately 50 days apart. The first peak in COVID-19 cases occurred following the first spike in PM_2.5_ on the 6^th^ July and the second peak of COVID-19 cases occurred after two spikes in PM_2.5_ in September. Simultaneous with this latter peak in PM_2.5_ there existed an ongoing wildfire called the “Bobcat fire” which started on 6^th^ September, and quickly spread after being fanned by irregular strong Santa Ana and Diablo winds. This wildfire was one of the largest in the history of wildfires in the vicinity of Los Angeles and may have contributed to the PM_2.5_ spike noted prior to the emergence of the B.1.429 variant (McKeever 2020). The persistent drought in South-West USA has further increased the PM_2.5_ levels (Borlina 2017).

A PM_2.5_ spike in the Washington Heights averaging 54.3+/-13.8µg/m3 (baseline 34.4(SD+/-11.6) was noted prior to the emergence of the B.1.526 variant was. This spike in PM_2.5_ in Washington Heights was noted in early September and from then on the new cases of COVID-19 progressively increased. A further rise in PM_2.5_ was noted in December coinciding with an increase in the positive gradient in COVID-19 new case diagnoses (Chart 11.). The main sources of outdoor air pollution in New York are motor vehicles and biomass combustion especially from wildfires. A recent study showed that pollutants from the smoke of wildfires from as far as Canada and the South-East U.S.A. caused significant increases in pollution concentrations in Connecticut and New York City (Rodgers et al 2020). Most of the reactive components from biomass burning are usually chemically transformed in the vicinity of the wildfire, however PM_2.5_ has a longer half-life ranging from a few days to about a week (Rodgers et al 2020).

Prior to May 2021 the B.1.2 variant in Louisiana and New Mexico was briefly conspicuous only to be soon displaced by the more transmissible B.1.1.7 variant. The PM_2.5_ levels in Louisiana and New Mexico were very similar whereby a peak of PM_2.5_ was noted 45 days before the detection and surge of the B.1.2 variant and another peak just before the variant’s surge. The PM_2.5_ levels in Louisiana and New Mexico reported atmospheric PM_2.5_ levels of 75+/-27.8µg/m3 (baseline 43.3 SD+/-14.4) µg/m3 and 71.4+/-11.3µg/m3 (baseline 43.6 SD+/-12.4) µg/m3 respectively. The baseline PM_2.5_ following the B.1.1.7 emergence was significantly lower 27.8µg/m3 (SD+/-18.0) suggesting that this variant acts more favourably at lower levels of PM_2.5_ as opposed to other SARS-CoV-2 variants (Muscat Baron 2021a). An increase of COVID-19 new cases occurred following the PM_2.5_ in Louisiana. In New Mexico a spike of COVID-19 new cases occurred approximately 45 days after the PM_2.5_ spike. The R.1. variant was detected in Eastern Kentucky during an outbreak in a Nursing home facility in mid-March 2021. This too seems to have been preceded by a PM_2.5_ spike recording an average of 37.7+/-7µg/m3 with a background baseline of 28.5µg/m3 (SD+/-6.8).

Potential contributors to elevated PM_2.5_ levels and the consequent spread of SARS-CoV-2 in the U.S.A. were a number of holidays encouraging mass gatherings. These included Independence Day 4^th^ of July, Labor Day in early September and Thanksgiving Day in late November.

### PM_2.5_ and the Emergence of the B.1.1.248 variant in Brazil

In Sao Paolo Brazil, two peaks of PM_2.5_ were noted prior to the emergence of the B.1.1.248 variant in early December. The first peak of PM_2.5_ started in mid-September and another peak in PM_2.5_ occurred in mid-October 2021. The average PM_2.5_ during its peak in Sao Paolo was 107.4µg/m3 (SD+/-34.2) before B.1.1.248 variant was detected when the baseline PM_2.5_ was 48.3µg/m3 (SD+/-18). The main cause of air pollution in Sao Paolo appears to be vehicular exhaust (Kabuto et al 1990). This is on the increase due to mass urbanization, increasing the population density which in turn elevates vehicular usage. The elevated population density in itself significantly increases human to human contact encouraging further SARS-CoV-2 transmission. Biomass combustion by the population and wildfires also contribute to the PM_2.5_ atmospheric load.

Wildfires in south-east Brazil produce smoke that aggravates air pollution in major cities such as Sao Paulo. In a study on asthma in children the average dose of PM_2.5_ was 1.95□μg/kg.day (CI: 1.62 – 2.27) during the dry season and 0.32□μg/kg.day (CI: 0.29 – 0.34) with the arrival of rain. During the dry season, children and adolescents showed a toxicological risk to PM_2.5_ of 2.07□μg/kg.day (95% CI: 1.85 – 2 .30) (de Oliviera et al 2012).

Depending on the weather, long-range transport of wildfire smoke affects the air quality of Sao Paolo. Combustion of biomass produces increased quantities of low-lying pollution exacerbated in part, to the South Atlantic subtropical high pressure system. Transported over considerable distances from wildfires, this pollution further contributes to poor air quality and smog in Sao Paulo (Targino et al 2019).

### PM_2.5_ and the Emergence of the B.1.617.2 variant in India

A broad peak in PM_2.5_ was noted during the first 3 weeks in January in Nagpur India where the B.1.617 variant emerged (Chart 18.). Prior to the emergence of the B.1.617 variant in India, the PM_2.5_ peak in Nagpur averaged 166.8+/-10.8µg/m3 in January 2021, with a baseline of 123.2µg/m3 (SD+/-16.9). Between the 14^th^ January and the 27^th^ April 2021, an estimated 9.1 million pilgrims attended the religious Hindu pilgrimage and festival called the Kumbh Mela. The peaks in PM_2.5_ coincided with the gatherings throughout the Kumbh Mela Festival, however the size of crowds decreased significantly towards the end of April possibly due to the increasing incidence and mortality of COVID-19. The beginnings of the surge of new cases with the Indian variant occurred in Maharashtra state of which Nagpur is a city, at the end of February (Chart 19.) (Quadri and Padala 2021).

In a study by Singh et al 2021, amongst all cities assessed, Delhi was found to have the highest air pollution, followed by Kolkata, Mumbai, Hyderabad, and Chennai. A common pattern was noted in most of these cities except for Chennai, whereby the highest concentrations of PM_2.5_ occurred in the winter while the lowest levels of this pollutant were noted during the monsoon season. PM_2.5_ levels in the cities exceeded WHO safety cut-off levels for 50% and 33% of days annually except for Chennai. In New Delhi for more than 200 days in a year exceeded the WHO safety cut-off levels. Compared to the previous years a decrease has been noted and can be attributed to the recent policies and regulations implemented in Indian cities attenuating air pollution. PM_2.5_ levels are however still elevated requiring stricter compliance to the Indian National Clean Air Program to further accelerate the reduction of the pollution levels (Singh et al 2021).

### PM_2.5_ and the Angiotensin II Converting Enzyme Receptor (ACE-2)

The occurrence of PM_2.5_ spikes a few weeks prior to the emergence of SARS-CoV-2 variants may have primed the effected populations to be more susceptible to COVID-19. The point of entry of the SARS-CoV-2 virus is the Angiotensin II Converting Enzyme Receptor (ACE-2) which is commonly found on type-2 pneumocytes responsible for gaseous exchange and mucus producing goblet cells. Specifically the ACE-2 protein acts as the receptor for the attachment of the SARS-CoV-2 spike protein, consequently increasing the risk for infection as well as severity of the disease in humans.

Preclinical studies utilizing the murine model exposed to particulate matter impacted both the ACE-2 protein and TMPRSS-2 (transmembrane protease serine type 2) (Sagawa et al 2021). Both the ACE-2 protein and TMPRSS-2 are required for the entry of SARS-CoV-2 into respiratory host cells. Immunohistochemical assessments indicated that exposure to particulate matter increased the expression of ACE-2 protein and TMPRSS-2. Image cytometry demonstrated increased expression of ACE-2 protein and TMPRSS-2 specifically in the type-2 pneumocytes which are potential targets for SARS-CoV-2. (Sagawa et al 2021). In another murine model endowed with human ACE-2 receptors, the bronchial instillation of particulate matter significantly increased the expression of ACE-2 and TMPRSS-2 in the lungs (Zhu et al 2021). Furthermore, particulate matter exacerbated the pulmonary lesions caused by SARS-CoV-2 infection in this mouse model (Zhu et al 2021).

In humans, during the COVID-19 pandemic in Italy, Borro et al showed that in response to exposure to PM_2.5,_ bioinformatic analysis demonstrated increased DNA sequences encoding for the ACE-2 receptor. The bioinformatic analysis of the ACE-2 gene identified nine recognized nucleic acid sequences for the aryl hydrocarbon receptor. In the same study correlations were noted between PM_2.5_ levels and COVID-19 incidence (R = 0.67, *p* < 0.0001), the mortality rate (R = 0.65, *p* < 0.0001) and the case fatality rate (R = 0.7, *p* < 0.0001) (Borro et al 2020).

Pollution with both nitrous oxide and PM_2.5_ has been shown to increase the concentration of ACE receptors in the lung (Paital et al 2021). Both pollutants induce an inflammatory change in the respiratory epithelium. This may occur with both chronic and acute inflammation. In an effort to cleanse the bronchial tree from pollutants, the number of mucus-producing goblet cells increase, with a consequent increase in these cells’ ACE-2 receptors (Paital and Agrawal 2021).

The above suggest a close link between exposure to particulate matter and the increase in the viral points of cell entry, the ACE-2. The spikes in PM_2.5_ may have acted differentially according to the time before variant emergence. The spike in PM_2.5_ approximately 50 days before the variant emergence, may have instigated an inflammatory response with a consequent induction in ACE-2 receptors in the respiratory tract. The PM_2.5_ spike immediately prior to the emergence of the variant may have indicated heightened human to human contact increasing viral load, which in the presence of readily available points of entry, increased infection rates. In the presence of high infection rates and a higher viral load the possibility of viral mutation became a greater possibility.

### Particulate matter acting as a Vector for SARS-CoV-2 Transmission

The COVID-19 pandemic appears to have spread during March 2020, from Wuhan in China, then to Qom in Iran and soon later to the Lombardy region in Northern Italy. An environmental variable common to all these three cities is the presence of elevated atmospheric levels of particulate matter (Muscat Baron 2020b) (Muscat Baron 2020c). Atmospheric particulate matter in the Lombardy region showed that out of 34 RNA extractions for the genes E, N and RdRP coding for SARS-CoV-2, twenty detected one of these genes (Setti et al 2020a).

High transmissions rates of SARS-CoV-2 infection were evident from outset of the pandemic (Muscat Baron 2020a). At the pandemic’s peak, the highest R_0_ SARS-CoV-2 achieved was estimated at 5.7 (Sanche et al. 2020), significantly higher than the R_0_ 1.4-1.67 of the H1N1 2009 Influenza pandemic (Coburn et al. 2009) and 2.2 for SARS-CoV-1 (Liu et al 2003). The substantially elevated R_0_ compared to other epidemics suggested that besides human to human transmission, other variables including particulate matter may accelerate SARS-CoV-2 spread (Setti et al 2020b).

Setti et al. 2020, showed that COVID-19 spread in 110 Italian provinces correlated with atmospheric levels of both PM_2.5_ and PM10. It was suggested that SARS-CoV-2 transmission could be further hastened by particulate matter carriage beyond the social distance of two metres up to 10 metres (Setti et al 2020b). In a similar study, PM_2.5_ and PM10 atmospheric levels correlated with COVID-19 infection rates in Italy and reaffirmed the hypothesis that particulate matter may also act as a vector for SARS-CoV-2 transmission (Comunian et al 2020).

In a teaching hospital in Kuala Lumpur Malaysia, a study showed that the highest SARS-CoV-2 RNA on PM_2.5_ in the ward correlated with number of patients with COVID-19 and the absence of air purifiers. High levels (74□±□117.1 copies μL−1) of SARS-CoV-2 genes on PM_2.5_ were noted in the single room ward without an air purifier compared to a general ward with an air purifier (10□±□7.44 copies μL−1) (Nor et al 2021).

The link between airborne pollution and infectious disease is not a novel one. Measles has the highest R_0_ of all infectious diseases with an R_0_ of 18 and has been closely associated with airborne pollution. In China, “dust events” in the Gansu region, have been linked with increased incidence of measles (Ma et al 2017). In the Niger, greater atmospheric pollution during the dry season is associated with measles-related childhood deaths which subside at the onset of the rainy season (Ferrari et al 2008). In 1935 during the Dust Bowl period, the state of Kansas experienced the most severe measles epidemic in USA history (Brown et al 1935). Following World War II, outbreaks of polio in the USA initiated at the beginning of summers and declined with the arrival of September rains (Oshinisky 2014).

### Mutational and Genotoxic Potential of PM_2.5_ on SARS-CoV-2

In January 2020 the genome of SARS-CoV-2 was sequenced indicating that it consisted of a single strand RNA containing approximately 29,000 nucleotide bases. Following the first sequenced RNA strand, a multitude of SARS-CoV-2 genomes have been sampled confirming that over 13,000 mutations had occurred in 2020 (Callaway 2020). The vast majority of mutants do not survive, however if the mutation confers an adaptation advantage, then through natural selection it will survive to outpace the incumbent variant.

Mutational potential on SARS-CoV-2 may occur through selective pressure due to an adaptation to the environmental milieu which may possibly be either mutagenic or genotoxic. The environmental factor may also come into play as a vector which may differentially favour mutants increasing their transmission. One such environmental factor may include airborne particulate matter. If this hypothesis is proven, it would be of singular importance as it may suggest that particulate matter may act as both a vector and mutagen for SARS-CoV-2. Moreover particulate matter may furthermore differentially favour variants by not only acting as a mutagen resulting in variant emergence, but actually be genotoxic to the progenitor virus expediting the latter’s disappearance.

It has been suggested that particulate matter PM_2.5_ acting as a vector, was not only responsible for SARS-CoV-2 transmission but may also have been involved in this virus’ evolution (Muscat Baron 2021a). Acting as SARS-CoV-2’s vector, the quantity and quality of PM_2.5_ may have exerted selective pressure determining the emergence of the first highly transmissible variant G614 in China (Muscat Baron 2021a).

A recurring set of mutations, namely E484K, N501Y, and K417N may suggest that SARS-CoV-2 is undergoing convergent evolution (Gupta 2021). This convergent evolution of SARS-CoV-2 may be due to the presence of a common mutagenic catalyst and vector in the form of the omnipresent pollutant PM_2.5_ which has repeatedly been shown to be a consistent co-factor in the SAR-CoV-2 Pandemic (Zoran et al. 2020)(Copat et al. 2020)(Sharma et al. 2020).

Contrasting effects by PM_2.5_ on viral infectivity have been shown possibly due to mutations in viruses and bacteriophages. Viral transmission may be considered a phenotypic reflection of genomic mutation. During the 1968 influenza pandemic and its aftermath, mutations were noted in the influenza virus hemagglutinin serotype H3 molecule and appear to have provided an advantage to the resultant variant to evade antibodies and consequently cause disease in previously immune individuals (Bean et al. 1992).

RNA viruses unlike DNA viruses lack the proofreading function of polymerase enzymes. This may be due to the hypothesized evolutionary precedence of RNA, which appears to have emerged well before the inception of DNA (Becker et al 2018). As opposed to DNA, RNA may act both as a genetic carrier and as an enzyme. RNA viruses have mutation rates resulting in 10^−3^ to 10^−4^ errors per incorporated nucleotide, which is significantly higher than DNA viruses’ error rate calculated at 10^−8^ to 10^−11^ errors per incorporated nucleotide (Alberts et al 2002). SARS-CoV-2 variants’ increased transmissible behaviour may therefore be dictated by these relatively frequent (compared to DNA viruses) RNA viral mutations (Fleischmann et al 1996). Viral mutagenicity due to particulate matter PM_2.5_ has been suggested with an experimental study on bacteriophage transmissibility in the presence of this pollutant (Groulx et al 2019). A study suggested differential effects on the transmissibility of two bacteriophages Φ6 and ΦX174 in the presence of PM_2.5._ Whereas the aerosol admixture of PM_2.5_ with Φ6 reduced this bacteriophage’s transmissibility, a diametrically opposite effect was noted on the bacteriophage ΦX174, demonstrating superior infectivity compared to controls (Groulx et al 2019).

### Adverse effects of Particulate Matter on Pulmonary Immunity

Particulate matter adversely affects pulmonary immunity at all levels of its defences. This is more so with PM_2.5_ as its micrometre diameters allow easy passage through the narrowest bronchioles and enter the alveoli. The mucociliary system is impaired by particulate matter disturbing its cleansing function of the respiratory tract. Pulmonary exposure to particulate matter appears to cause mucociliary paresis and promotes goblet cell mucus hypersecretion similar to tobacco smoking (Xu et al. 2020) (Muscat Baron 2020d).

The respiratory epithelium is rendered more permeable at the cell junctions and cell membranes due to particulate matter. Respiratory epithelial permeability to viral invasion is further encouraged by allowing particulate matter-induced pro-inflammatory mediators which weaken the baso-lateral aspect of respiratory host cells and reducing the concentration of tight junction proteins (Liu, et al 2019).

Particulate matter may encourage uncontrolled viral dissemination by disturbing macrophage function. Inefficient clearance by viral phagocytosis may be due to deficient Human leucocyte antigen recognition, inhibition of cytokine production and altered function of toll-like receptor genes expressed on cell membranes (Fujimoto et al. 2000). Particulate matter exposure may influence ability of Natural Killer cells to eliminate cells infected by viruses (Müller et al. 2013). Exposure to particulate matter PM_2.5_ was linked to increased levels of antiangiogenic and proinflammatory cytokines (Pope et al. 2016).

Particulate matter exposure appears to affect CD4+ and CD8+ T cells leading to the suppression of interleukin IL-2 and interferon 1-γ production (Pierdominici et al. 2014). In severe cases of SARS-CoV-2 infection due to resultant significant loss of CD4+ and CD8+ T cells early in the infection, there appears to be delayed adaptive immune responses, leading to prolonged viral clearance (Channappanavar, et al 2014).

### Increased Infectiousness and Antibody Escape due to SARS-CoV-2 Variants – a mutagenic role of Particulate Matter?

Evidence is emerging that SARS-CoV-2 variants have increased transmissibility and could evade neutralization by antibody responses elicited by previous infection and vaccines. Serum samples from patients who recovered from COVID-19 or received a vaccine could not completely neutralize new variants such as the South African B1.351 variant which possesses a mutation called E484K (Tegally et al 2020). Another mutation N105Y, is also linked to weakening antibody activity against the SARS-CoV-2 (Andreano et al 2020). There are also the K417N and K417T mutations which in concert with the combination of the E484K and the N105Y mutations appear to accelerate the lethality of (Khan et al 2021).

Transmissibility and antibody evasion are the hallmarks of increased infectiousness. More comprehensive studies (Wang et al. 2021) have shown that most observed mutations strengthen the binding between the spike protein’s receptor binding domain and the ACE-2 receptor. More recently found mutations of the receptor binding domain, such as the N439K, S477N, S477R, and N501T, further cement viral RBD and ACE-2 binding, increasing infectiousness.

Wang et al (2021) carried out genetic and protein-protein binding analysis elucidating that antibody evasion may be due to a number of mutations in the spike protein’s receptor binding domain. Besides the E484K and the N501Y mutation involved in United Kingdom (UK), South Africa, and Brazil variants many more variants have been found to weaken the binding between the receptor binding domain and neutralizing antibodies. Further disruption of receptor binding domain linkage with neutralizing antibodies was noted with the mutations L452R and E484Q found in the Indian variants. The L452R mutation is found in the California variant B.1.427.

Receptor binding domain mutations may pose a threat to vaccine escape, threatening the efficacy the current vaccines available. A list of most likely vaccine escape mutations include S494P, Q493L, K417N, F490S, F486L, R403K, E484K, L452R, K417T, F490L, E484Q, and A475S. The mutation T478K appears to make the Mexico B.1.1.222 the most infectious variant Wang et al (2021) and this may account for the high case-fatality ratio noted in this country (John Hopkins Resource Centre).

Wang et al suggested that the genetic evolution of the SARS-CoV-2 receptor binding domain may be due to a combination of four factors. These include host cell gene editing, viral proof reading and random genetic drift with natural selection over-arching all these three factors (Wang et al. 2021). It would be interesting to explore whether the presence of airborne particulate matter in elevated levels, as noted in the spikes of this pollutant preceding the variants’ emergence, may actually catalyze these three factors, introducing the environmental element of natural selection. The impact of particulate matter on the propagation of the ACE-2 receptor has already been confirmed. This may set a chain reaction to affect both host cell gene editing and viral proof reading and instigate the catalyzed genetic drift.

### Spikes in PM_2.5_ prior to the Emergence of SARs-CoV-2 Variants-An Ecological Fallacy or a Surrogate for increasing Viral Burden

There is literature that has not confirmed the role of particulate matter in the seeding and spread of COVID-19. A study done by Ong et al. 2020, demonstrated that SARS-CoV-2 could not be detected in all of the air samples assessed. A caveat to this study indicated that the short sampling time of ¼ hr–4 h might not be representative of the total air volume in the ward and the presence of SARS-CoV-2 might have possibly been diluted during air exchanges in the ward (Ong, S. W. et al. 2020).

Another study in Northern Italy by Collovignarelli et al. put the association between particulate matter and COVID-19 in doubt. Collovignarelli et al. excluded a significant correlation between atmospheric particulate matter and the incidence of COVID-19. This latter study suggested that there may be other factors, including meteorological factors,that may have synergised with particulate matter to spread COVID-19. If one were to look closely at the provinces in Northern Italy as regards seeding and doubling time of COVID-19, the highest rates are to be found in land-locked provinces especially in Lodi, Bergamo, and Aosta. The impact of the vicinity to the sea as a factor determining COVID-19 rates has been alluded to (Muscat Baron 2020b) (Muscat Baron 2020c). By virtue of the absence of particulate matter sources from the sea, the level of this pollutant is likely to be lower and diluted in coastal provinces. Moreover the increased sodium chloride content in particulate matter derived from marine sources may have had a role in deterring the adhesion of the hydrophobic regions in the SARS-CoV-2 spike protein to particulate matter (Muscat Baron 2020c).

One may surmise that the occurrence in the spikes of this pollutant in relation to the emergence of variants was just a coincidence. This is unlikely as assessing all the data available on the World Air Quality Index indicates two characteristic patterns of PM_2.5_ levels. In some cities such as Beijing and Sao Paolo, PM_2.5_ levels demonstrate perennial elevations in December and January as in the case of China or October in Brazil. The other pattern involves unique rises of atmospheric concentrations such as in Kent U.K. and Washington Heights New York.

On the other hand the association between particulate matter and COVID-19 has its support. The increase in pulmonary ACE-2 receptors and its effect on respiratory immunity after exposure to particulate matter has been scientifically confirmed (Paital and Agrawal, 2020), (Sagawa et al., 2021). The presence of SARS-CoV-2 genes on particulate matter provides circumstantial evidence of the potential vector effect of this airborne pollutant (Setti et al 2020b).

It is also biologically plausible that particulate matter’s mutagenic and genotoxic effect may also affect SARS-CoV-2 both outside and inside the host, leading to the emergence of variants and the displacement of the progenitor virus. The uncanny similarity in the patterns of PM_2.5,_ prior and during the emergence of the SARS-CoV-2 variant suggests a biological link. Lastly the appearance of similar mutations in the different variants, increasing their transmissibility, in widely disparate and distant regions, lends its itself to suggest that convergent evolution of SARS-CoV-2 is occurring in the presence of an ubiquitous environmental factor.

## Limitations of the Study

The average daily PM_2.5_ levels are taken in this study and it must be mentioned that there may be wide variations between the maximum and minimum levels of particulate matter throughout the day including when there may be human exposure to this pollutant. Exposure to atmospheric outdoor PM_2.5_ was carried out in this review, does not necessarily equate that to humans being similarly exposed to the same levels indoors. A collation of five studies demonstrated that indoor SARS-CoV-2 transmission was very high compared to outdoors (18.7 times; 95% confidence interval, 6.0–57.9) studies compared to outdoor spread (<10%) (Bulfone et al 2021). Although this study utilized outdoor PM_2.5_, the study by Brunekreef et al. mentioned earlier, demonstrated that indoor PM_2.5_ concentrations highly correlated with outdoor concentrations (median R= 0.7-0.8) (Brunekreef et al 2005). In some countries, such as in China, indoor levels may actually be higher if fossil fuels are used for heating purposes. In Wuhan in 2020, natural gas consumption for heating purposes increased 2.8 times fossil fuel utilization by the traditional stove and water heater, potentially producing another indoor source for particulate matter (Dong et al 2020).

There was a significant difference in new case counts obtained between different states included in this review. This suggests the strong possibility of variation in the frequency and availability of diagnostic testing for SARS-CoV-2. From the Beijing data available only the latter part of the bell-shaped curve for the new case counts could be assessed. These COVID-19 new cases in both Wuhan and Beijing were preceded by spike in PM_2.5_. In this study not all the states assessed had daily COVID-19 new case counts available.

The detection and emergence of variants has its drawbacks especially from the temporal aspect. There is a great variation in the availability and access to RNA sequencing processes. The USA (less than 1% of global COVID-19 sequencing) has the greatest capacity for sequencing but the absence of a single Health System as in the U.K. (9% of global sequencing) hampers the co-ordination of sampling and its availability for sequencing. There is also a significant cost to sequencing procedures (Maxmen 2021). A model has been proposed that within the SARS-CoV-2 perspective, 5% sampling of all positive tests in a population would allow the detection of emerging variants at a prevalence between 0.1% to 1.0%. Attenuating the risk of vaccine escape and the prevention of future coronavirus pandemics very much depends on the availability and easy access to genomic surveillance (Vavrek et al 2021).

## Conclusion

There appears to be a link between atmospheric PM_2.5_ and the emergence of SARS-CoV-2 variants. In most regions assessed, two groups of PM_2.5_ spikes were noted prior to the emergence of SARS-CoV-2 variants. These spikes in PM_2.5_ spike may suggest that the combination of a number of factors including, a).Anthropogenic activity increasing the viral burden, b). PM_2.5_-induced propagation of the ACE-2 receptor (the viral point of host cell entry), c). Potential PM_2.5_-induced viral mutagenesis resulting in variant emergence and genotoxicity to the progenitor, d). PM_2.5_ toxicity diminishing host pulmonary immunity and e). Possible PM_2.5_ vector effect, increased the prospect of the emergence SARS-CoV-2 variant. The above findings suggest that significant changes in PM_2.5_ levels may not only contribute to transmission, but also to the evolution of SARS-CoV-2.

N.B. At the time of submitting this paper the Delta Plus Variant (Nepal variant) became evident as variant of interest. Preliminary evaluation indicated that this variant was detected at the end of January 2021 and elevated PM_2.5_ levels were noted in Katmandu at the middle and end of December 2020 and extremely high levels at the beginning of January 2021.

## Data Availability

Availability of all data from links below

https://aqicn.org

https://waqi.info

https://coronavirus.jhu.edu/data

---

**Chart 1.**
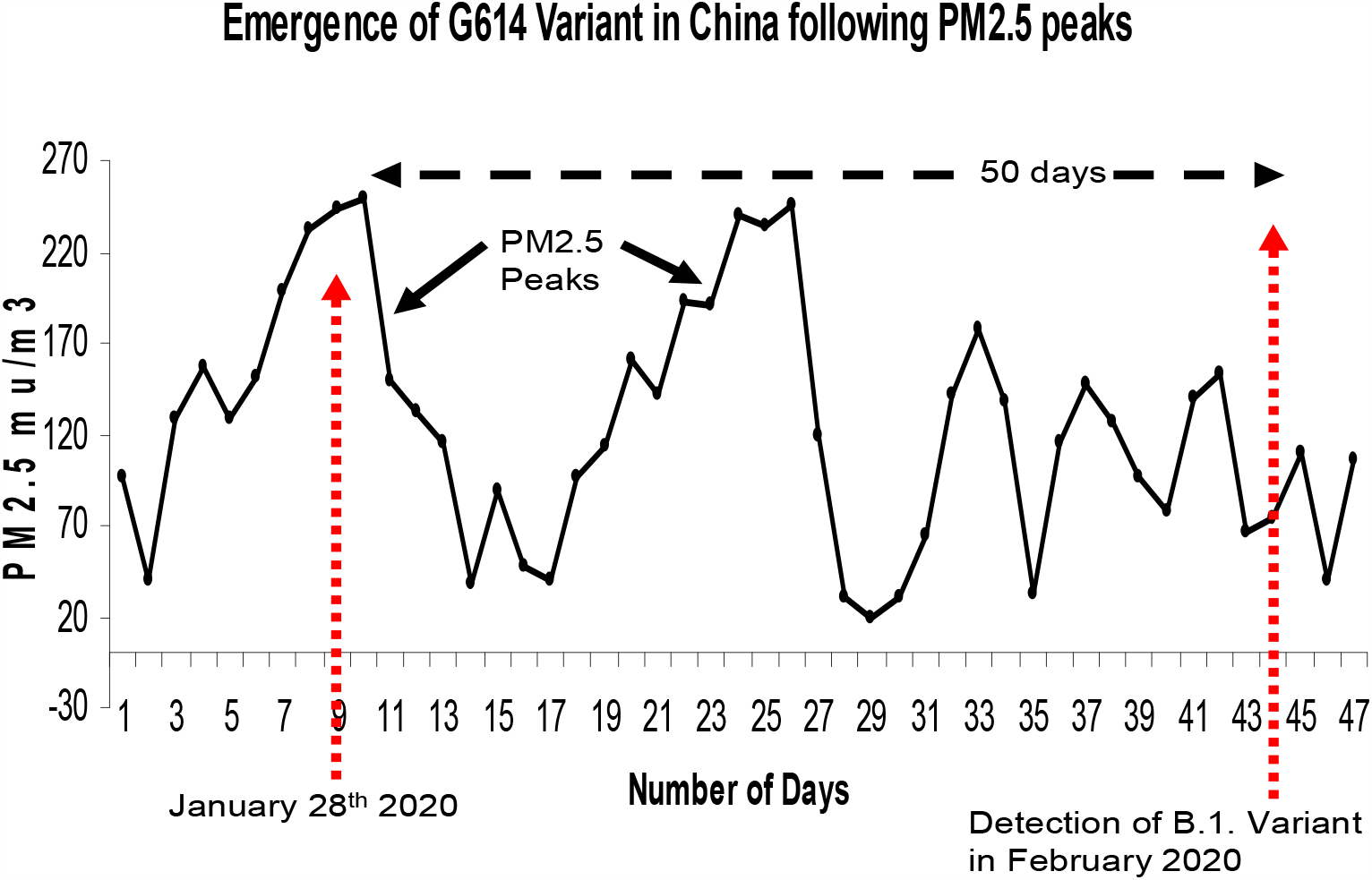
Emergence of G614 Variant in Beijing after atmospheric PM_2.5_ spikes in the mid-January 2020.

**Chart 2.**
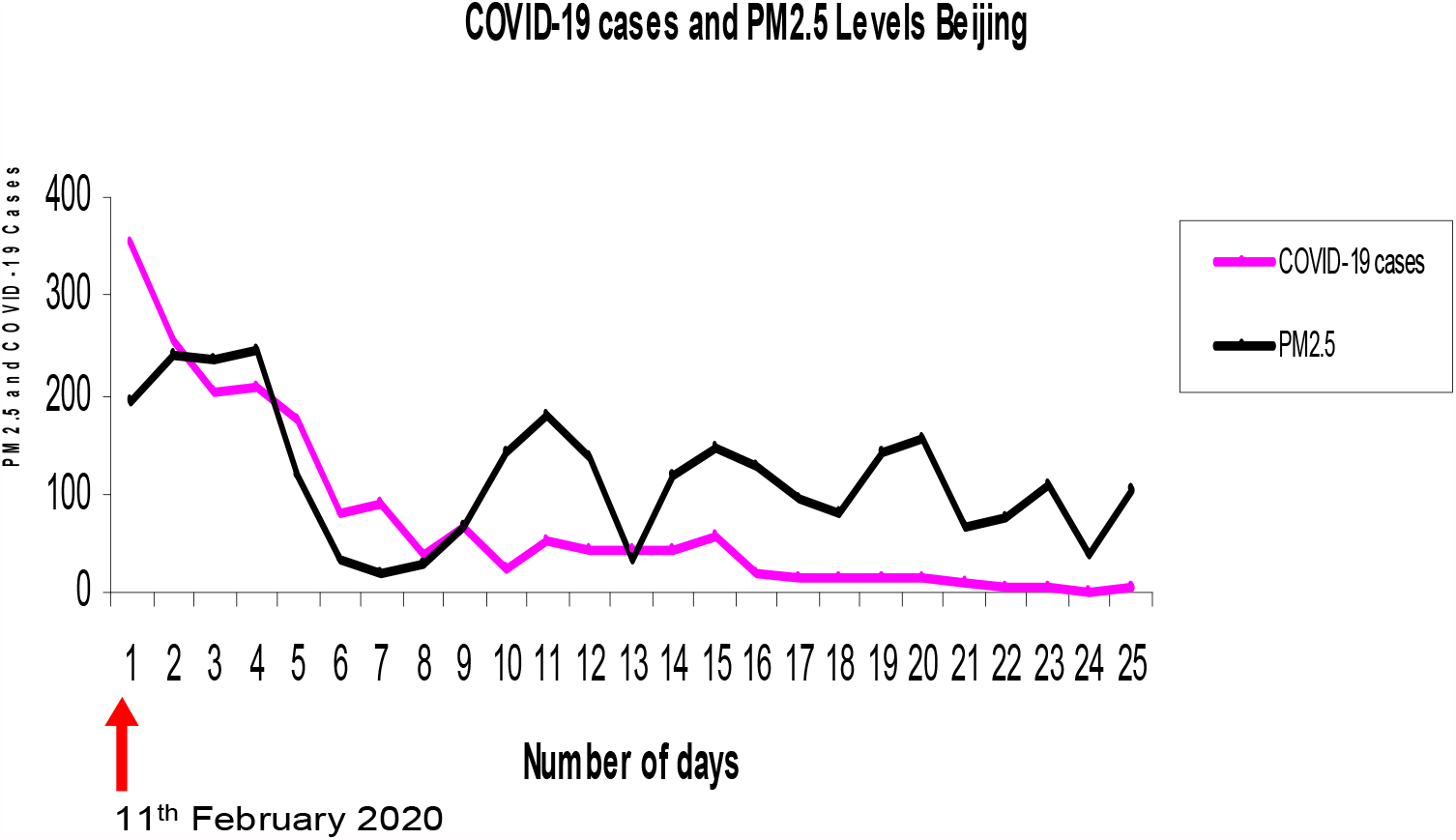
Emergence of G614 Variant and New cases of SARS-CoV-2 in Beijing after atmospheric PM_2.5_ spikes in the mid-Febuary 2020.

**Chart 3.**
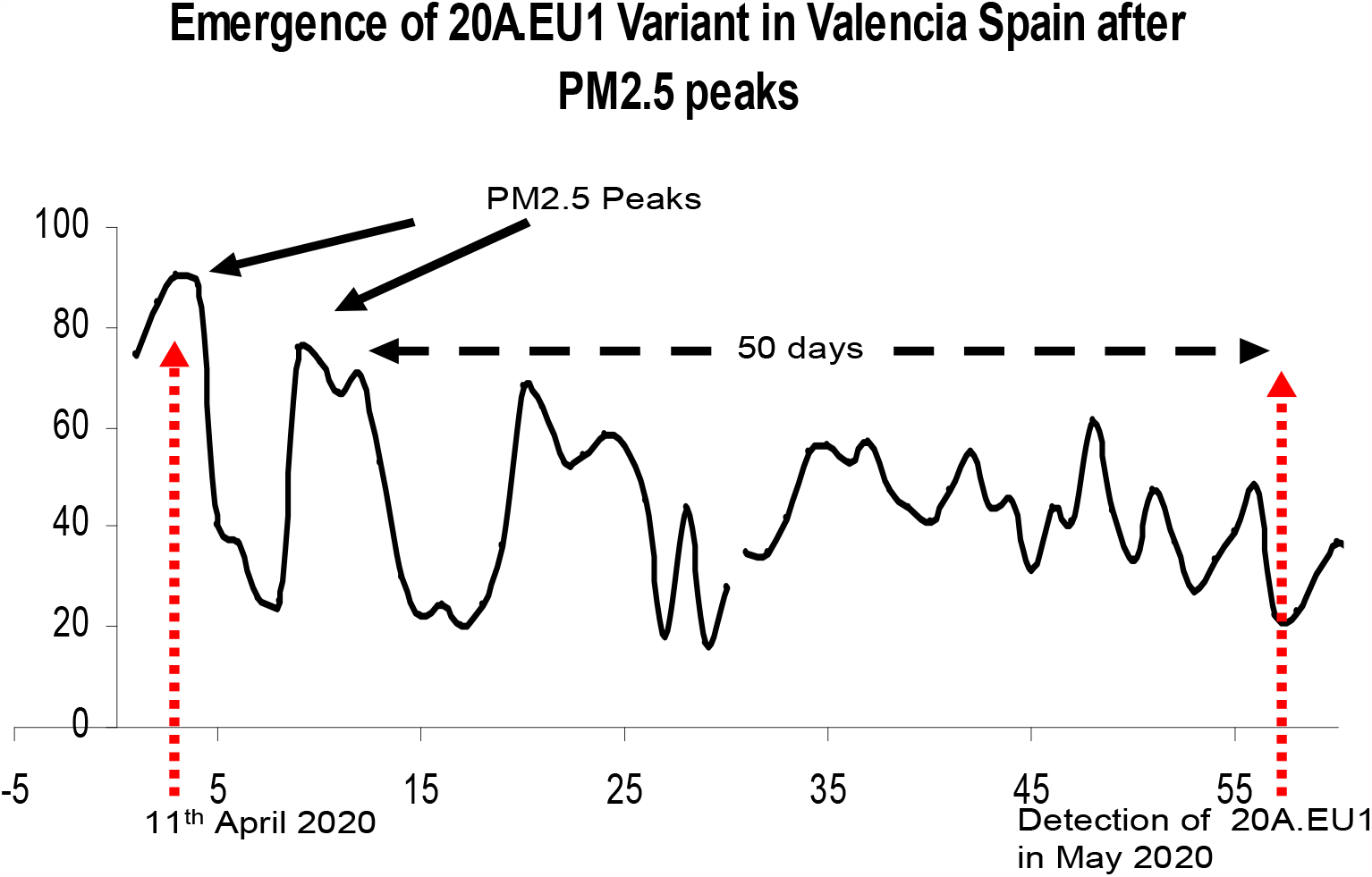
Emergence of 20A.EU1 Variant in Valencia Spain after atmospheric PM_2.5_ spikes in the mid-April 2020.

**Chart 4.**
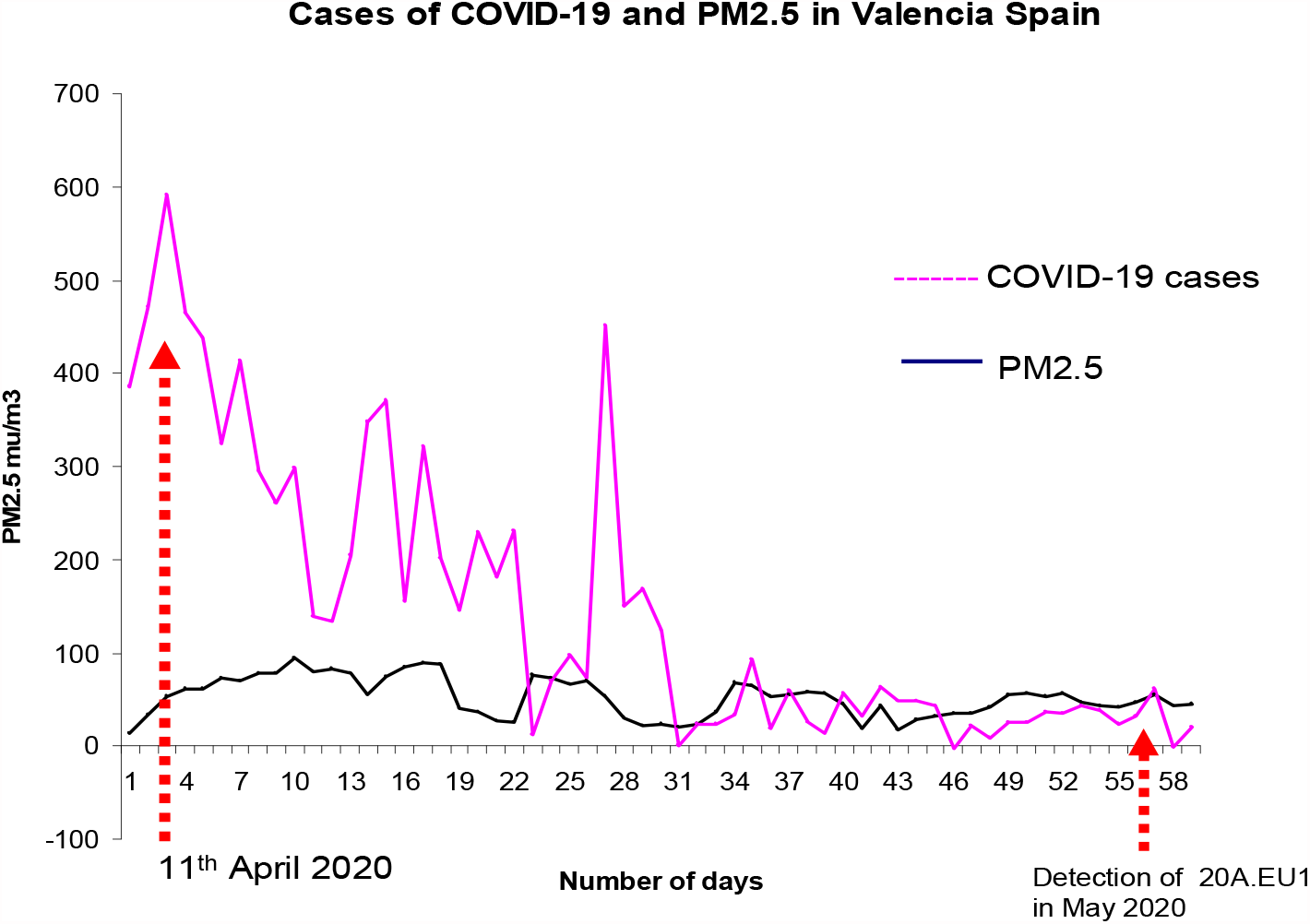
Emergence of 20A.EU1 Variant and New Cases of COVID-19 in Valencia Spain after atmospheric PM_2.5_ spikes in the mid-April 2020.

**Chart 5.**
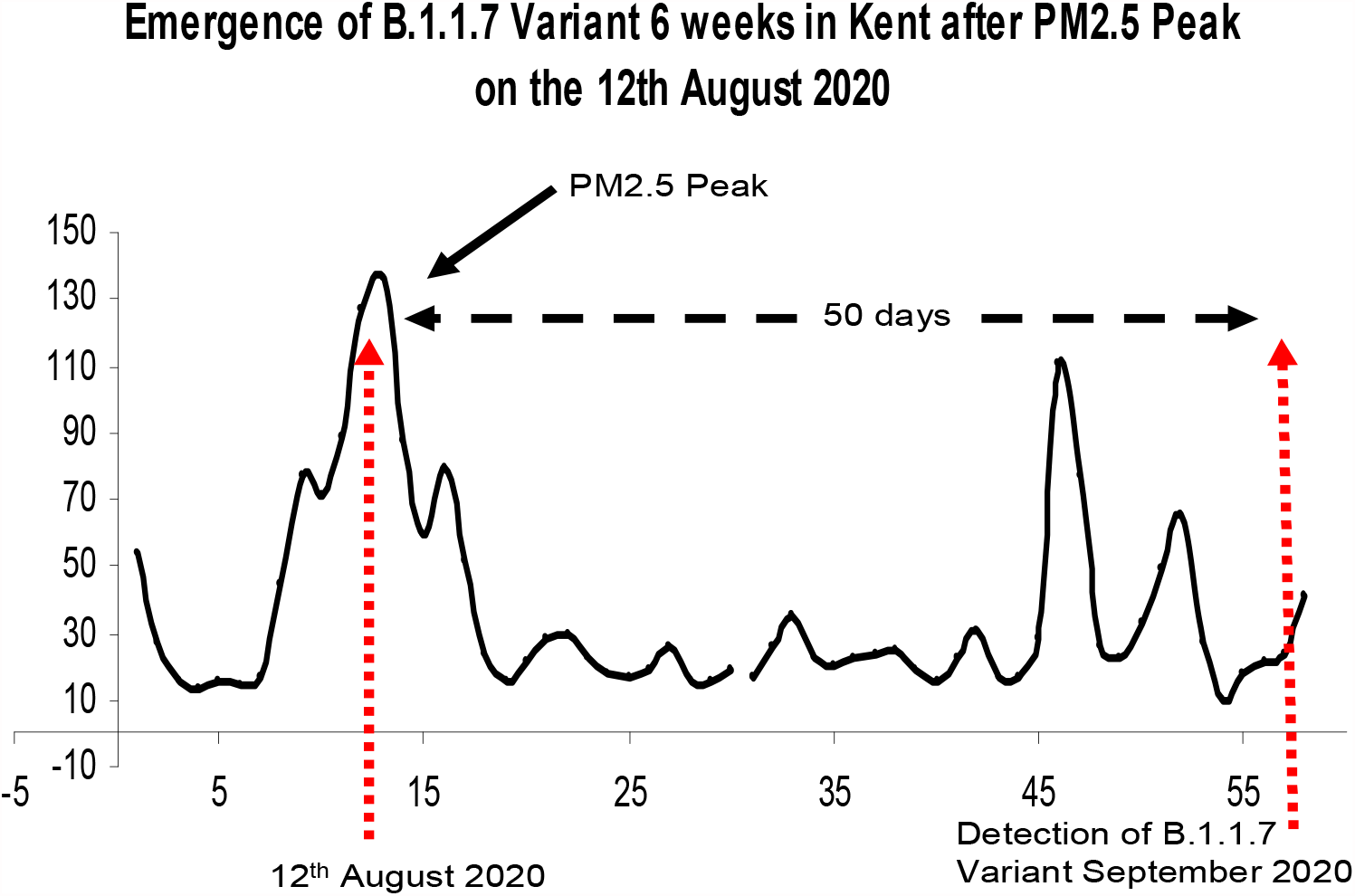
Emergence of B1.1.7 Variant in Bexely U.K. after spikes of atmospheric PM_2.5_.

**Chart 6.**
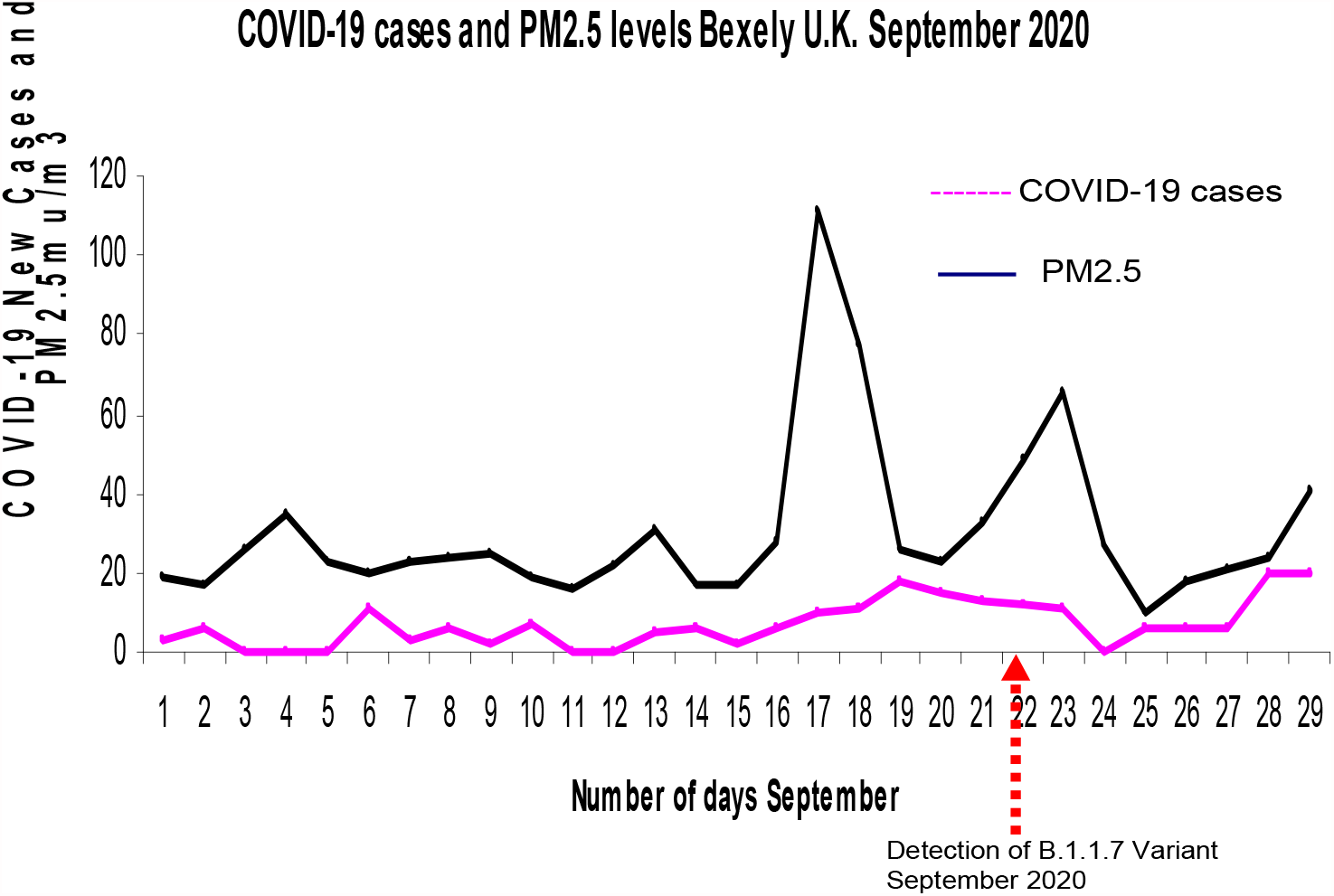
Emergence of B1.1.7 Variant and New Cases of COVID-19 in Bexely U.K. after spikes of atmospheric PM_2.5_ in September 2020.

**Chart 7.**
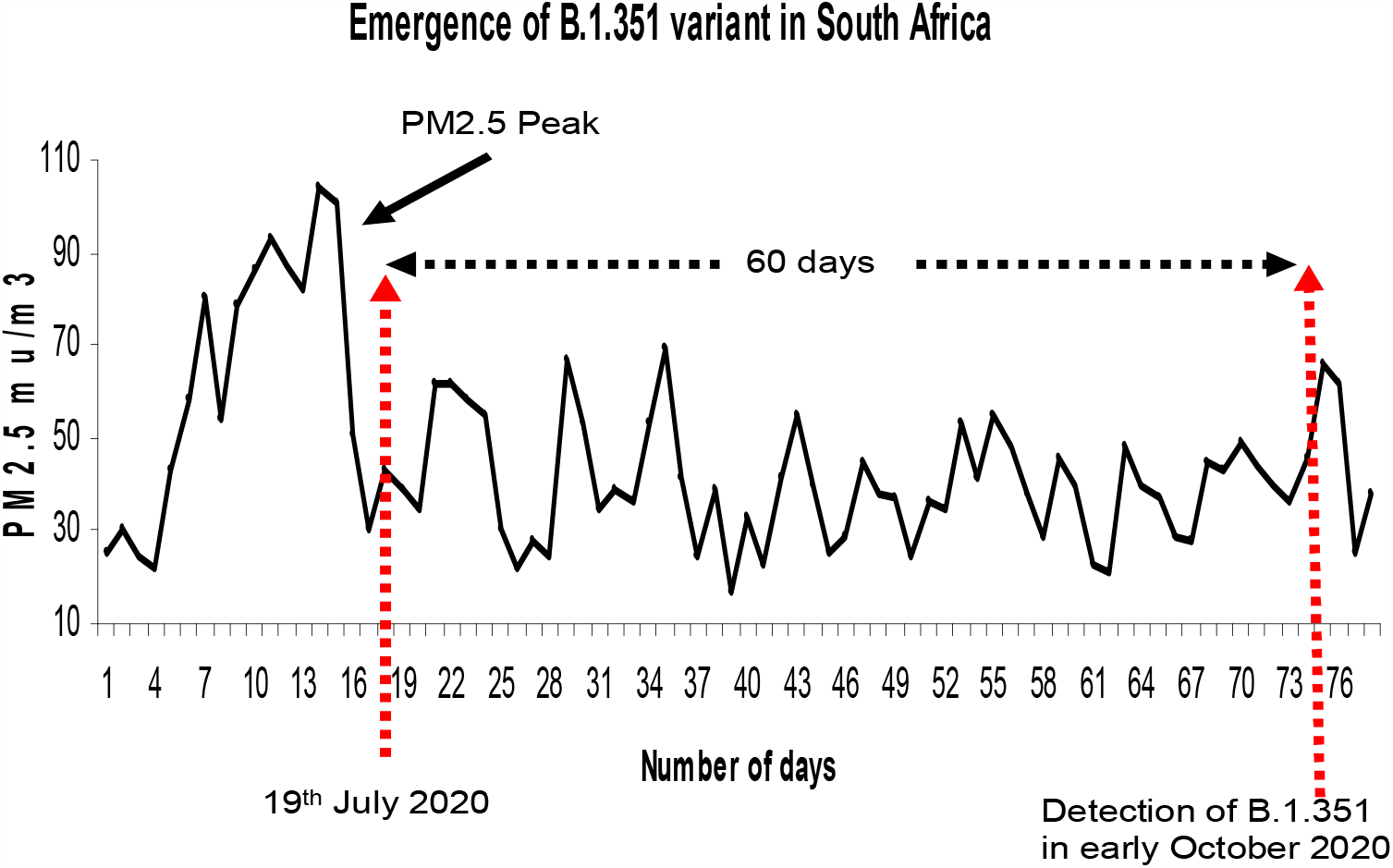
Emergence of B1.351 Variants in South Africa after spikes of atmospheric PM_2.5_.

**Chart 8.**
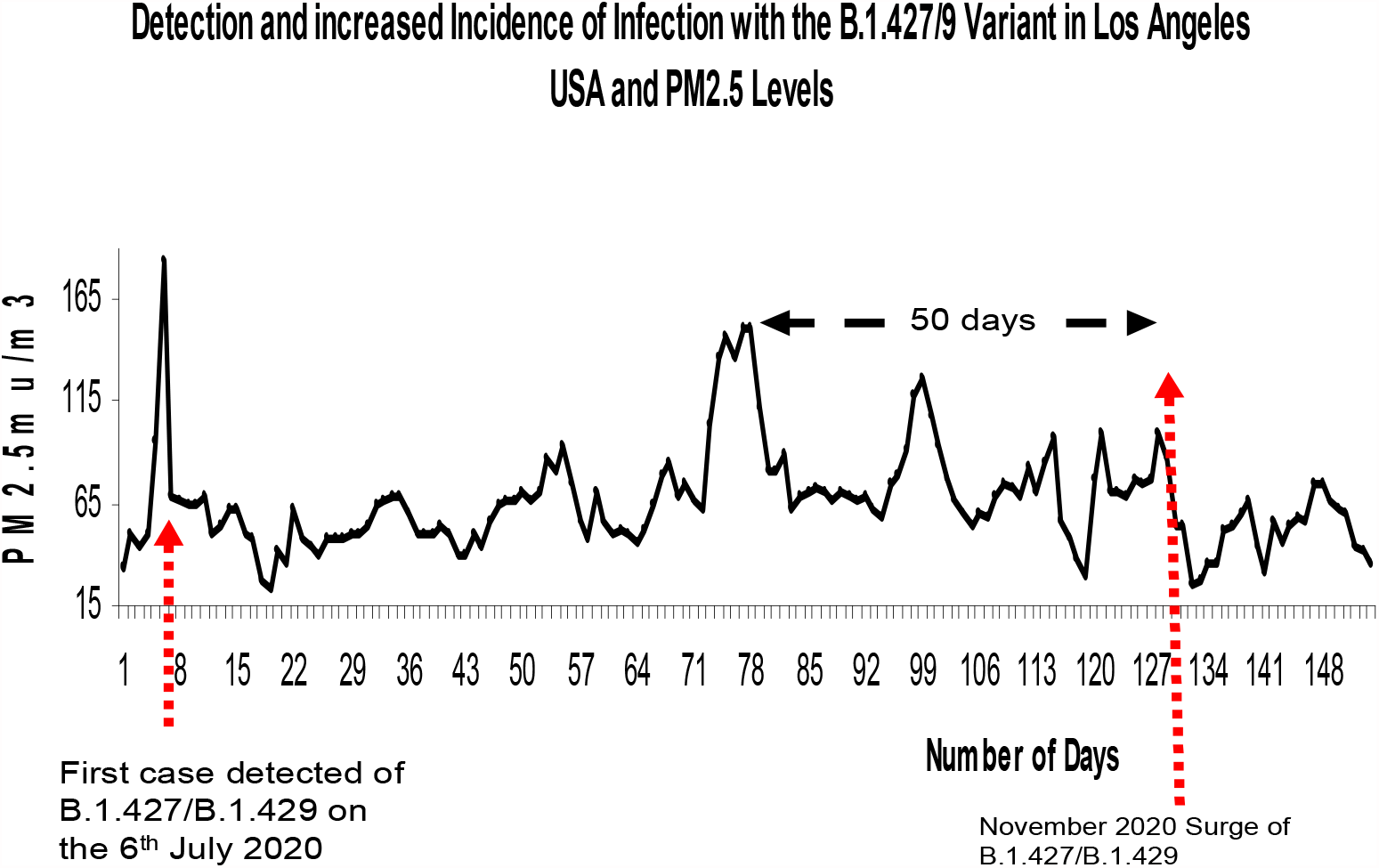
Emergence of B1.429 Variants Los Angeles after spikes of atmospheric PM_2.5_.

**Chart 9.**
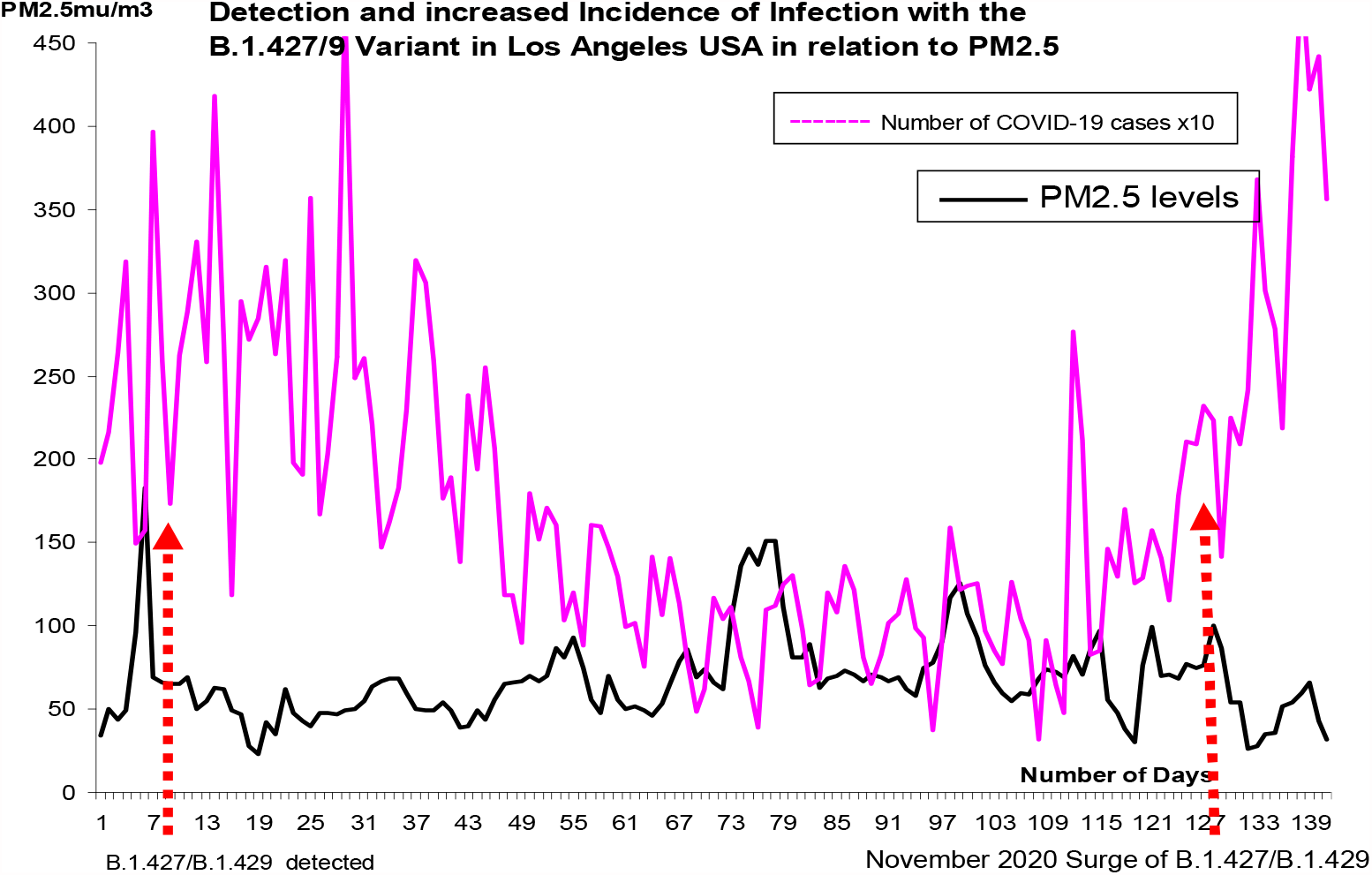
Emergence of B1.429 Variants and New cases of COVID-19 Los Angeles after spikes of atmospheric PM_2.5_.

**Chart 10.**
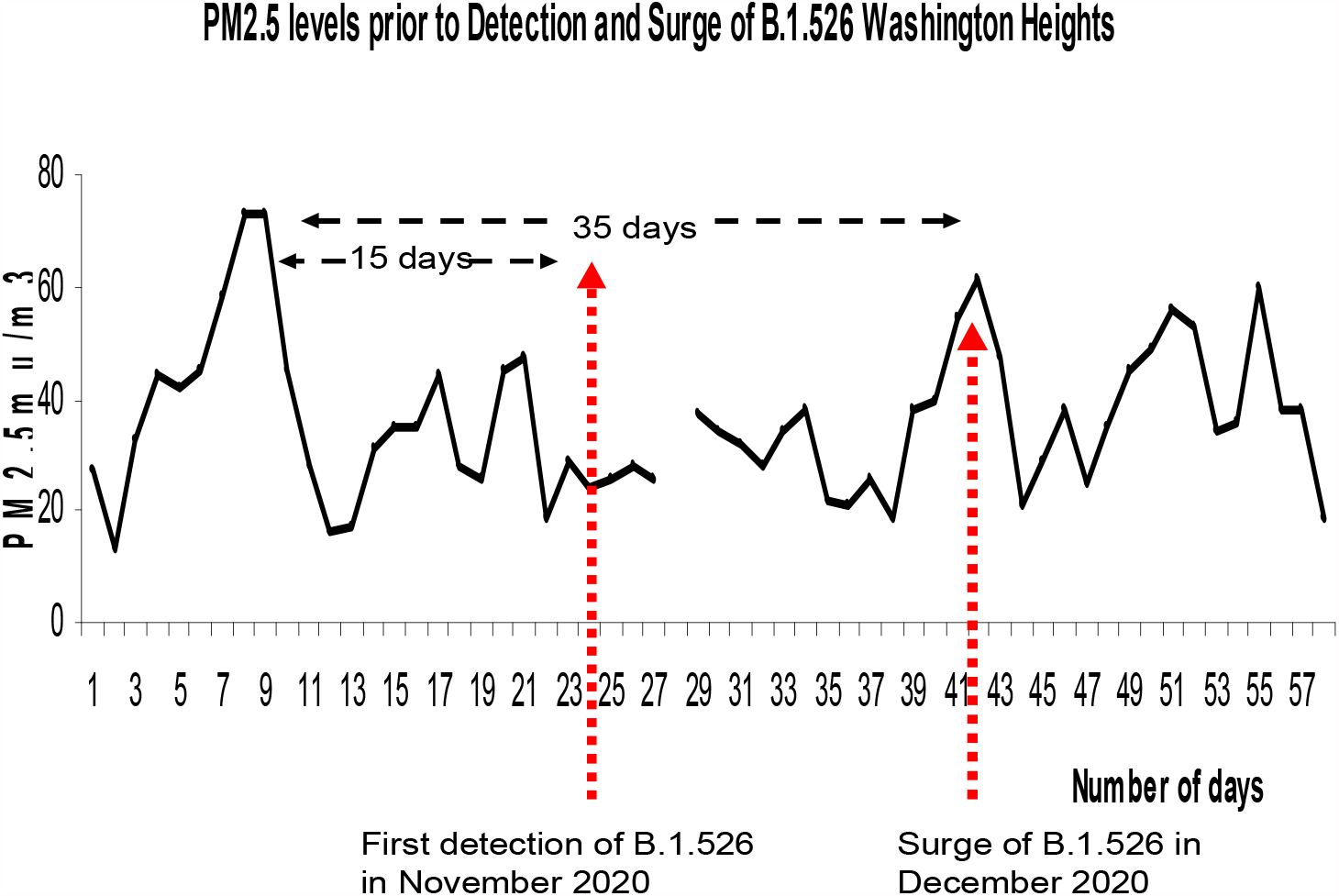
Emergence of B1.526 Variants in Washington Heights New York after spikes of atmospheric PM_2.5_.

**Chart 11.**
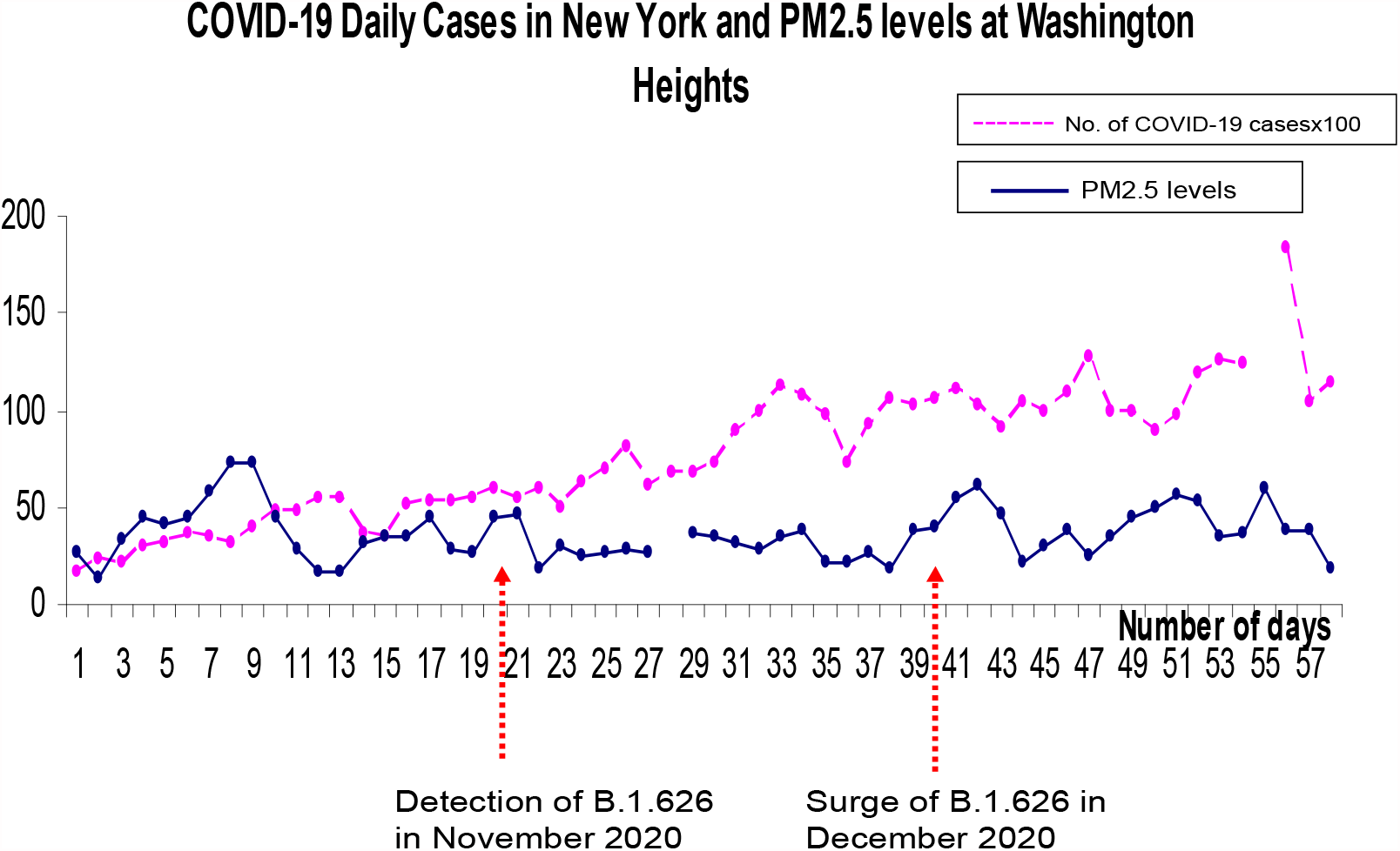
Emergence of B1.526 Variants and New cases of COVID-19 in Washington Heights, New York after spikes of atmospheric PM_2.5_.

**Chart 12.**
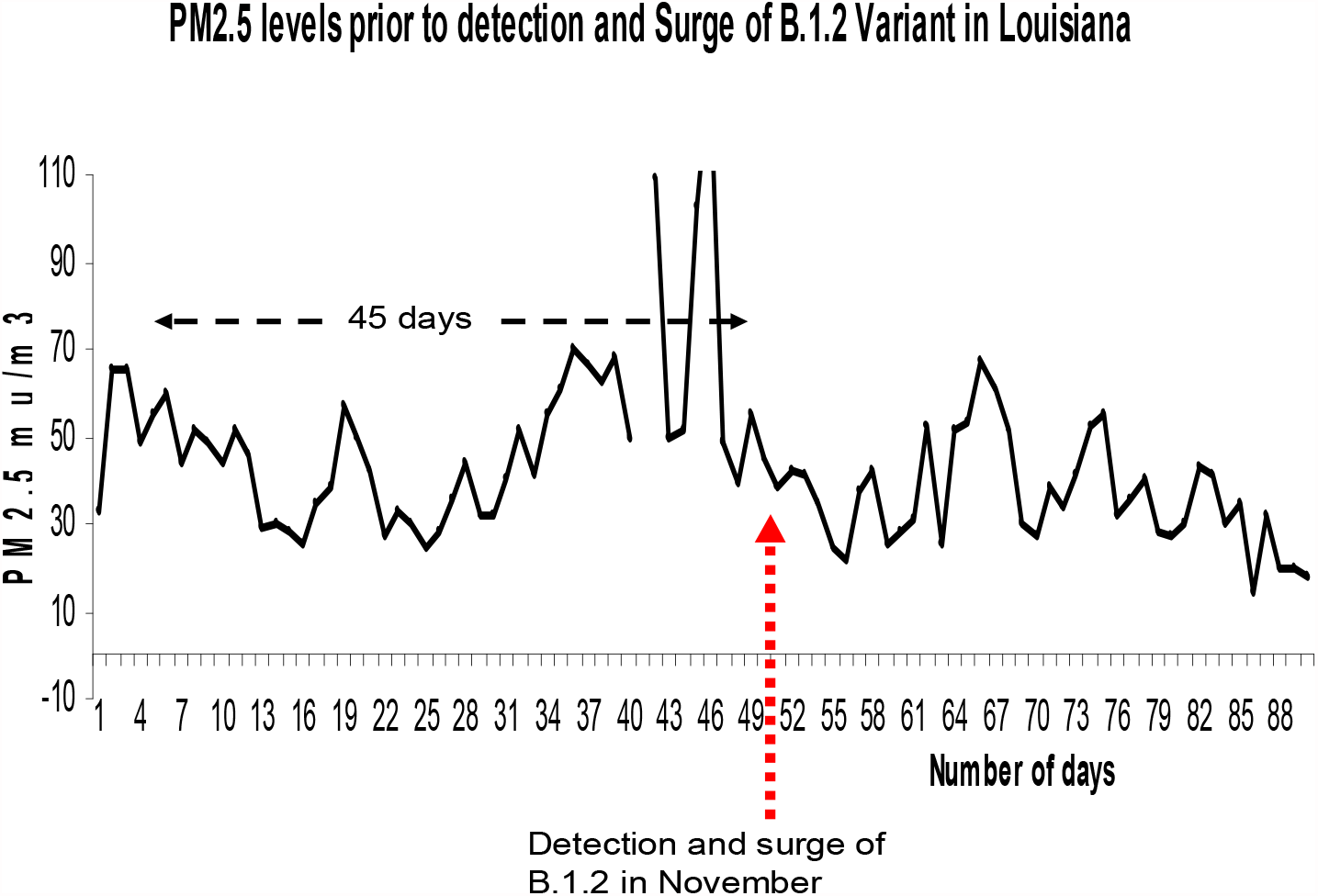
Emergence of B1.526 Variants in Louisiana USA after spikes of atmospheric PM_2.5_.

**Chart 13.**
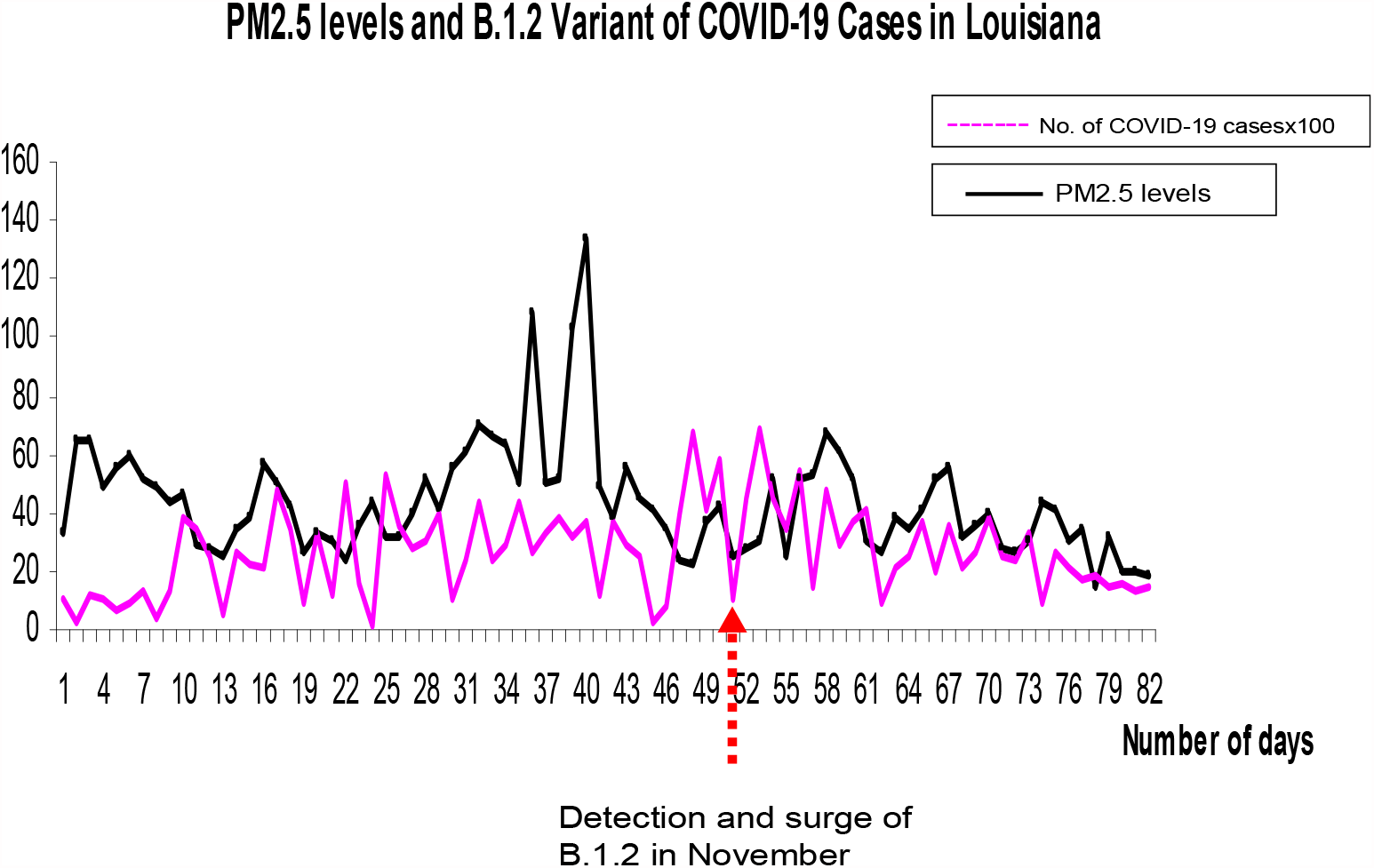
Emergence of B1.2 Variants and New cases of COVID-19 in Louisiana, USA after spikes of atmospheric PM_2.5_.

**Chart 14.**
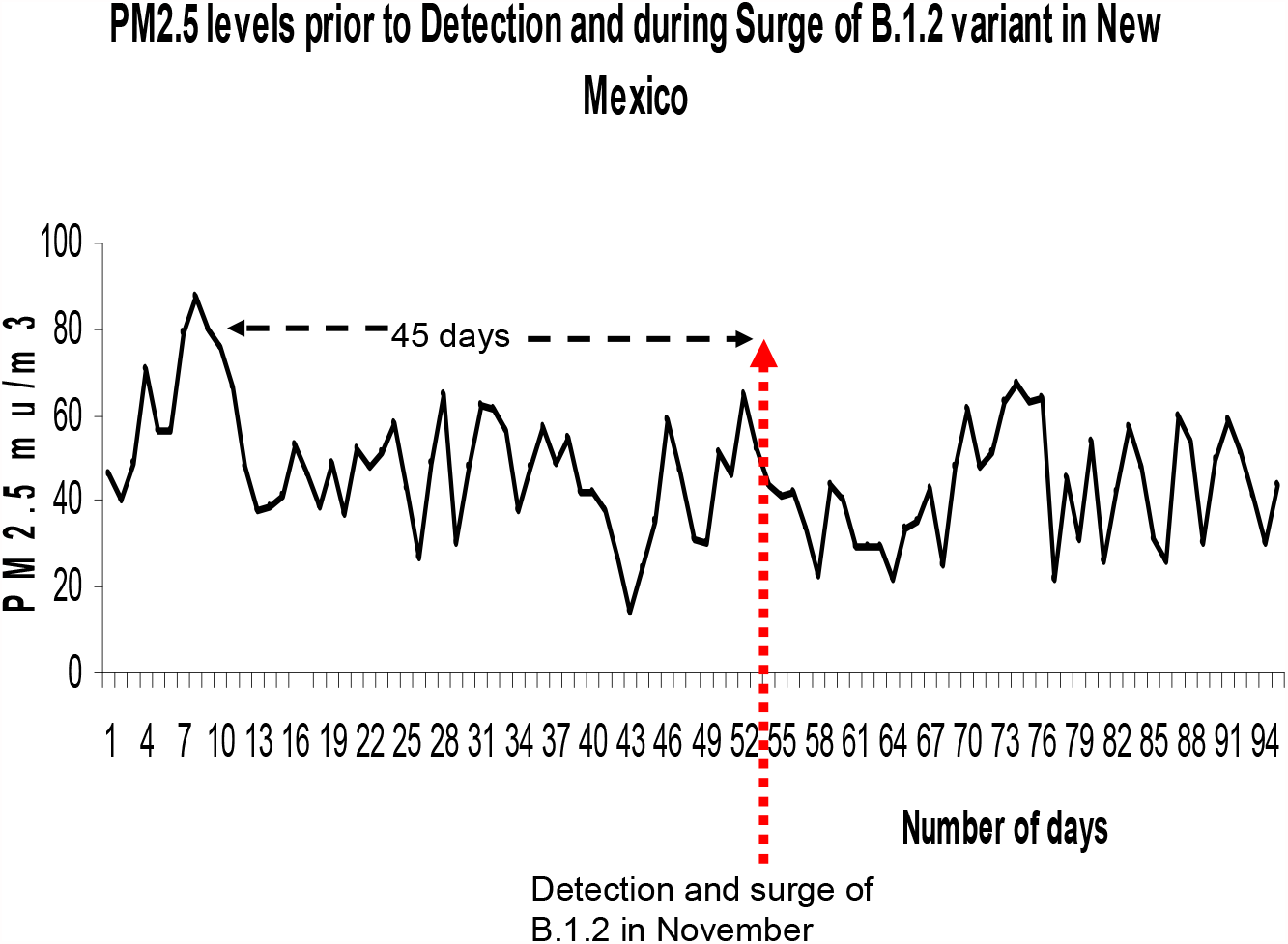
Emergence of B.1.2 Variants in New Mexico, USA after spikes of atmospheric PM_2.5_.

**Chart 15.**
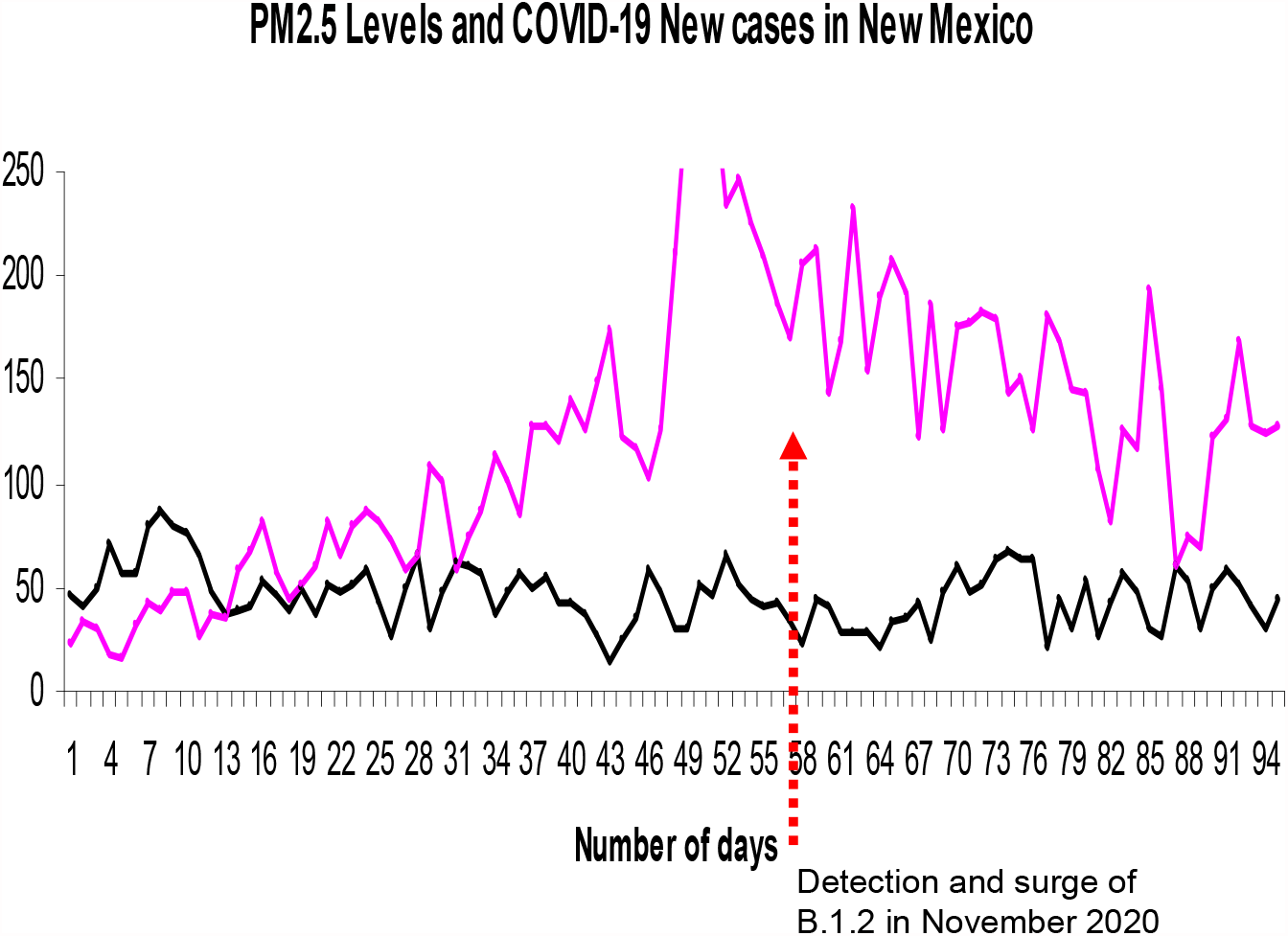
Emergence of B.1.2 Variants in New Mexico, USA after spikes of atmospheric PM_2.5_.

**Chart 16.**
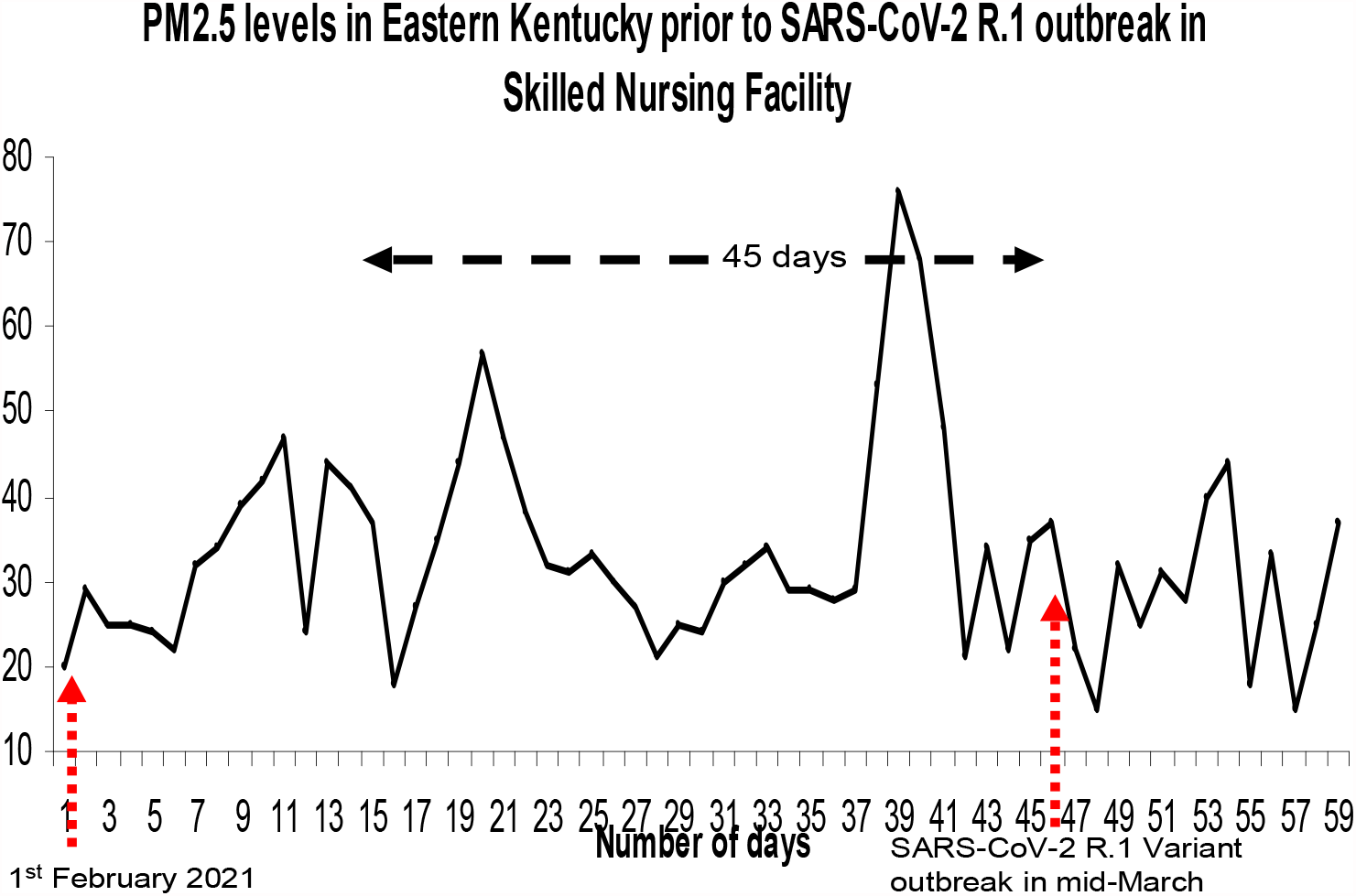
Emergence of R.1. Variant in Kentucky, USA after spikes of atmospheric PM_2.5_.

**Chart 17.**
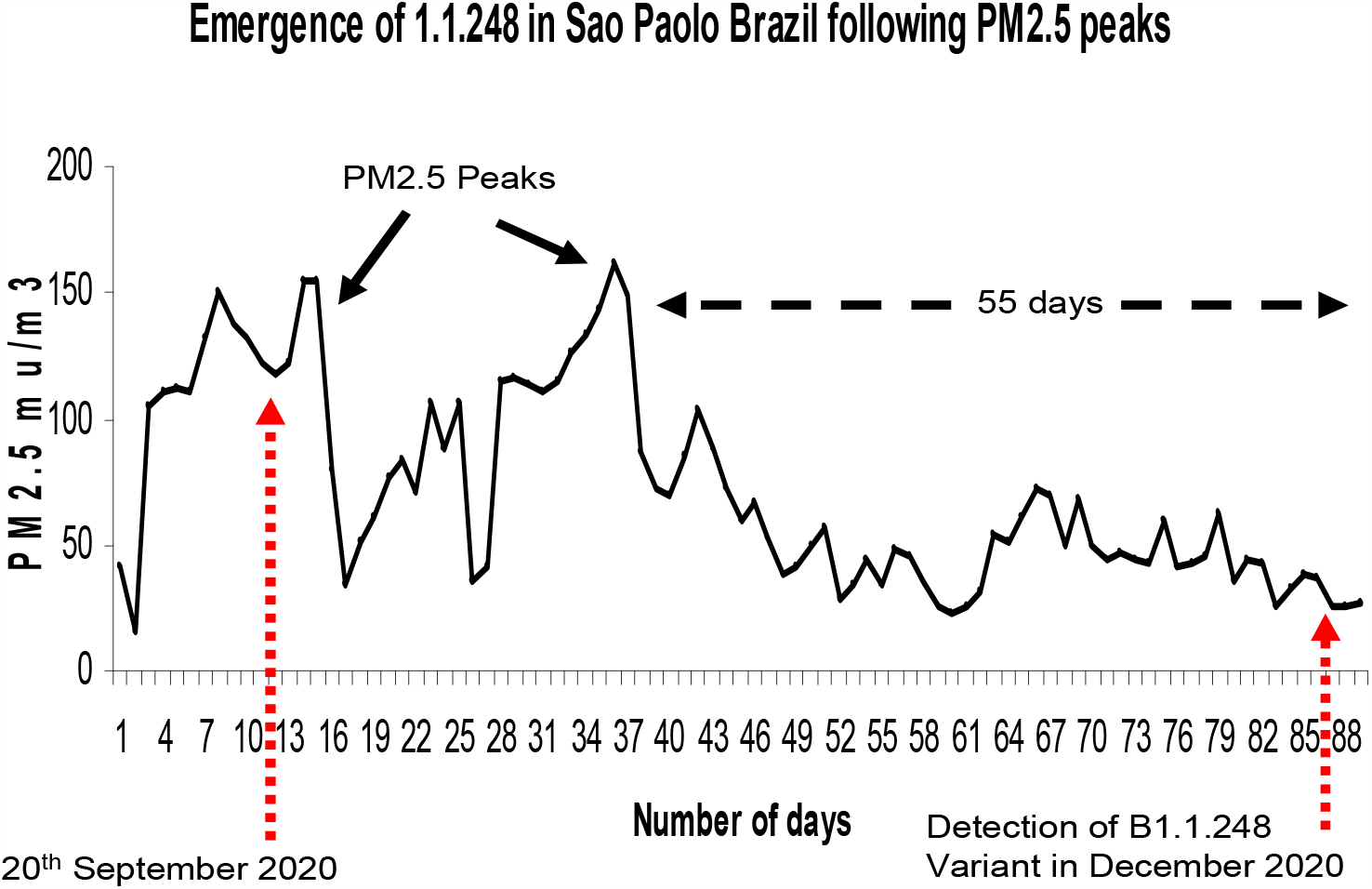
Emergence of B1.1.248 Variant in Brazil after spikes of atmospheric PM_2.5_.

**Chart 18.**
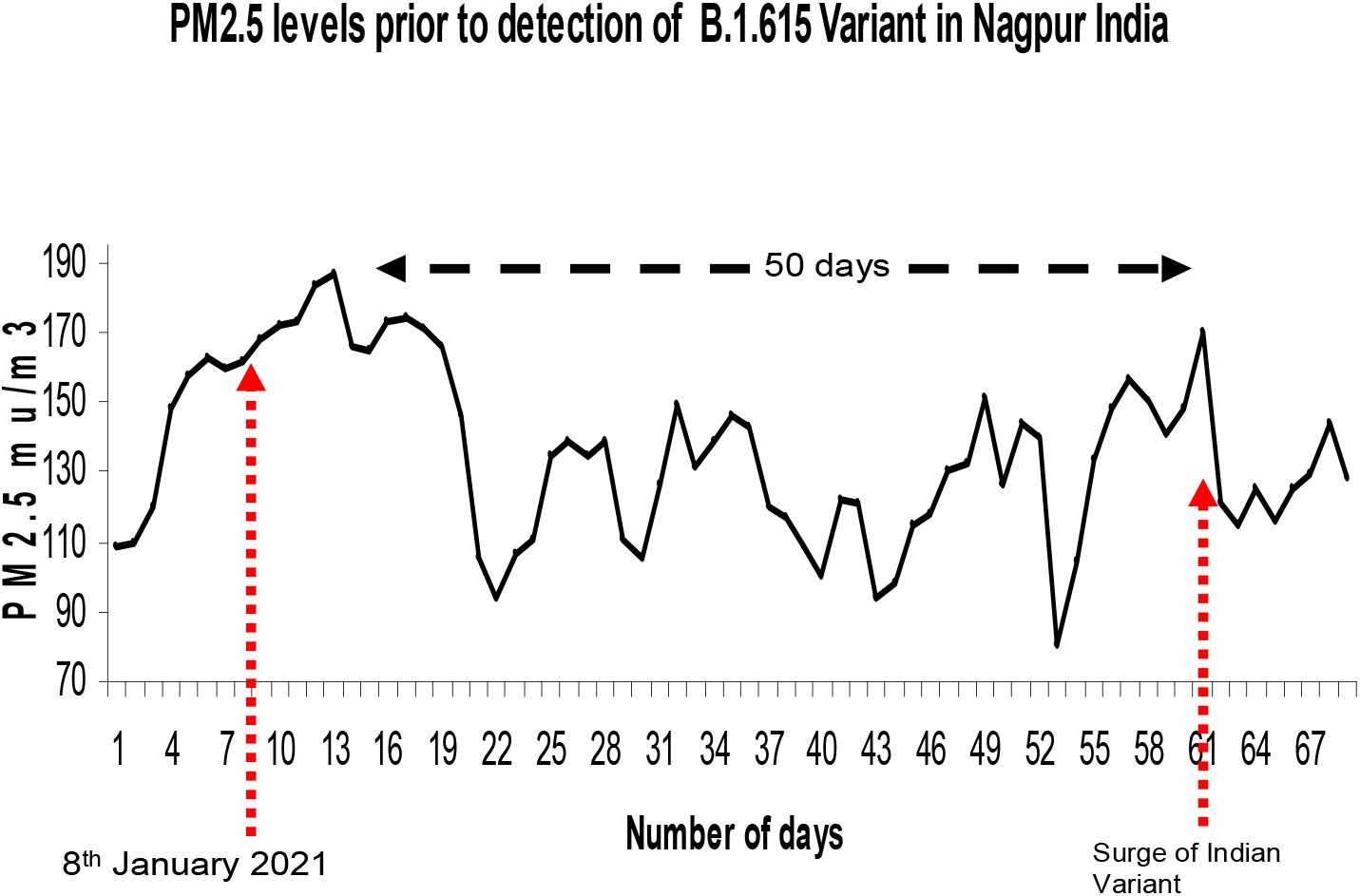
Emergence of B1.617 Variant in Nagpur India after spikes of atmospheric PM_2.5_.

**Chart 19.**
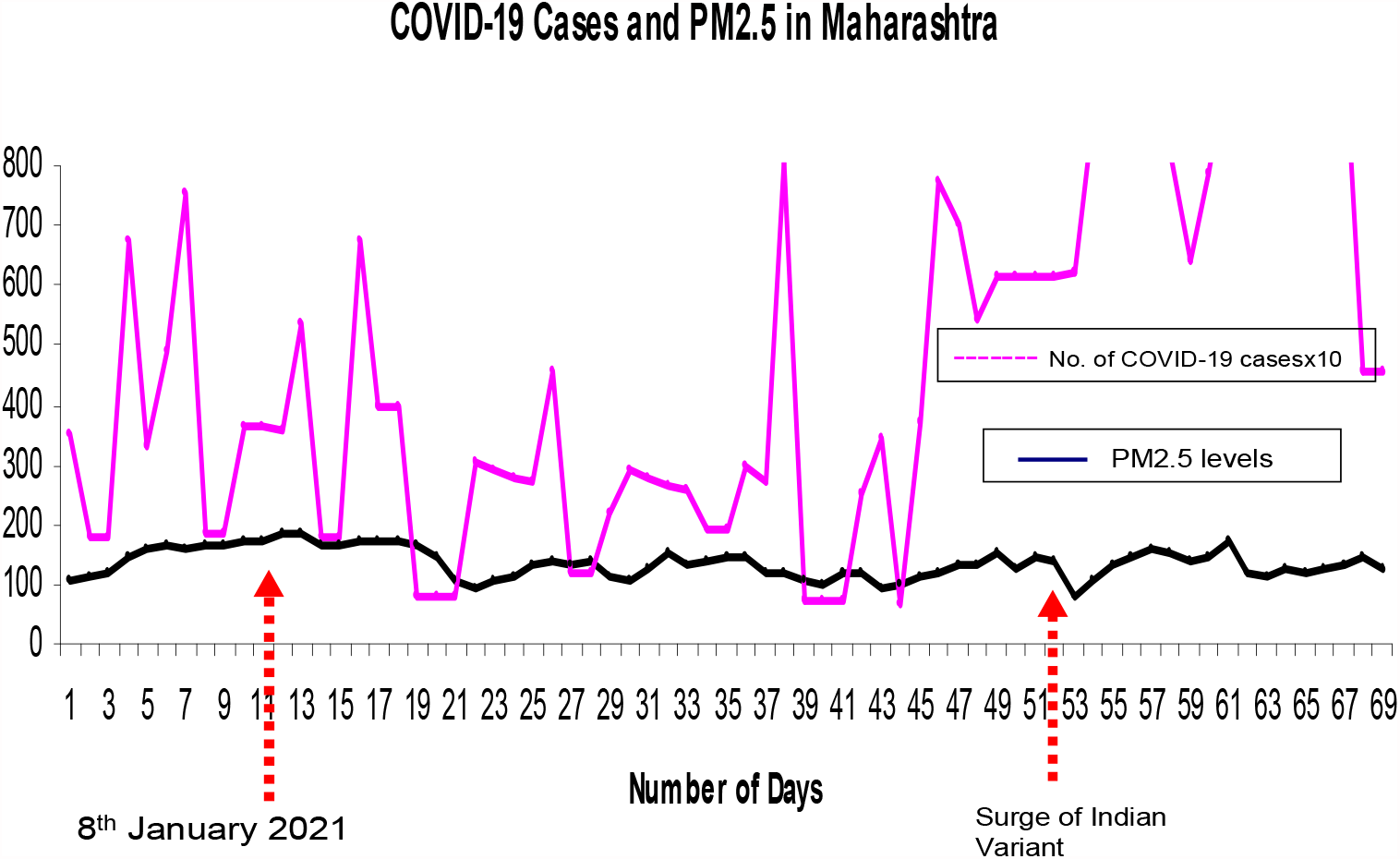
Emergence of B1.617 Variant and New Cases of COVID-19 in Marahashtra, India after spikes of atmospheric PM_2.5_ in Nagpur.

**Chart 20.**
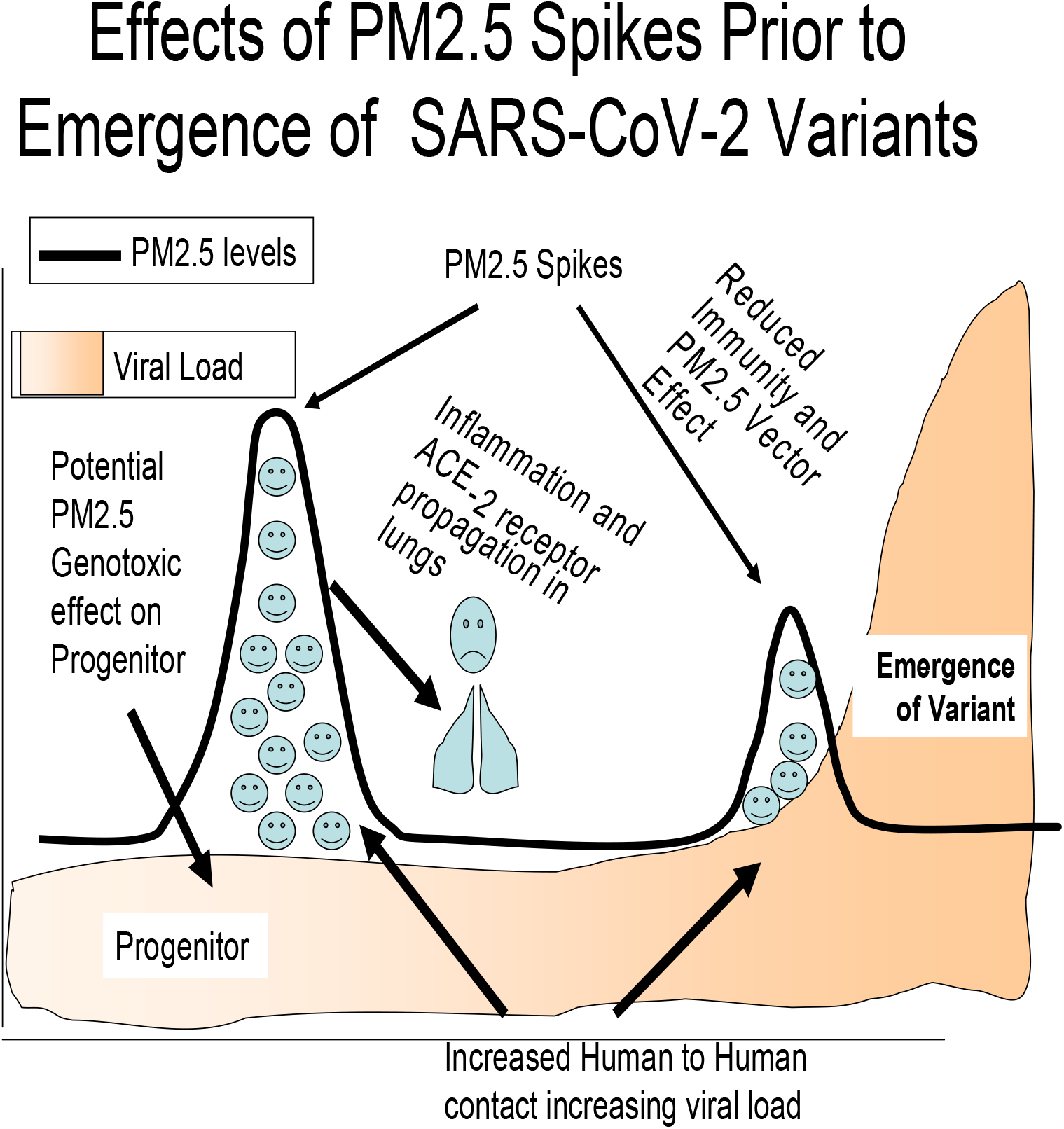
Schematic view of Atmospheric PM_2.5_ levels with contemporaneous viral load leading to Emergence of SARS-CoV-2 Variant. First PM_2.5_ spike associated with anthropogenic effect due to increased human activity and contact, leading to lung inflammation, ACE-2 receptor propagation, and potential PM_2.5_ viral mutagenic induced variant emergence and genotoxicity to the progenitor expediting the latter’s eventual displacement by the emerging variant. Second PM_2.5_ spike also related to diminished population immunity and potential PM2.5 vector effect.

## Notes

### Competing Interest Statement

The authors have declared no competing interest.

### Clinical Trial

N/A

### Funding Statement

No Funding

